# RT-RPA-Cas12a-based discrimination of SARS-CoV-2 variants of concern

**DOI:** 10.1101/2022.05.11.22274884

**Authors:** Guiyue Tang, Zilong Zhang, Wei Tan, Fei Long, Jingxian Sun, Yingying Li, Siwei Zou, Yujiao Yang, Kezhu Cai, Shenwei Li, Zhiyi Wang, Jiakun Liu, Guobing Mao, Yingxin Ma, Guo-Ping Zhao, Zhen-Gan Tian, Wei Zhao

**Author notes:** These authors contributed equally (Guiyue Tang, Zilong Zhang, Wei Tan).

## Abstract

Timely and accurate detection of SARS-CoV-2 variants of concern (VOCs) is urgently needed for pandemic surveillance and control. However, current methods are limited by the low sensitivity, long turn-around time or high cost. Here, we report a nucleic acid testing-based method aiming to detect and discriminate SARS-CoV-2 VOCs by combining <u>R</u>T-<u>R</u>PA and <u>C</u>RISPR-Cas12a <u>d</u>etecting assays (RRCd). With a detection limit of 10 copies RNA/reaction, RRCd was validated in 204 clinical samples, showing 99% positive predictive agreement and 100% negative predictive agreement, respectively. Critically, using specific crRNAs, representatives of single nucleotide polymorphisms and small deletions in SARS-CoV-2 VOCs including N501Y, T478K and ΔH69-V70 were discriminated by RRCd, demonstrating 100% accuracy in clinical samples with *C*_t_ < 33. The method completes within 65 min and could offer visible results without using any electrical devices, which may facilitate point-of-care testing of SARS-CoV-2 and its variants.

## Introduction

The pandemic of COVID-19 has caused 494.5 million infections and 6.1 million deaths as of April 10^th^ 2022 (https://covid19.who.int/table). The pathogen of this pandemic, SARS-CoV-2, has undergone rapid and constant mutations in the process of transmissions, bringing worldwide uncertainty to the diagnostics, vaccine effectiveness and therapeutics (Tao et al. 2021).

Since November 2020, generally five rapidly expanding SARS-CoV-2 lineages have been discovered and designated as variants of concern (VOCs), *i*.*e*., the Alpha, Beta, Gamma, Delta, and Omicron (Oude Munnink et al. 2021) (**Supplementary Figure 1**). Besides the current Omicron, among these lineages, the Alpha and Delta VOCs were dominant in frequency over a relatively long period of time. With gradual resumption of air travel, Customs, and ports, certain previously isolated areas were riskily exposed to the SARS-CoV-2 variants. These variants, either with higher infectivity or increased pathogenicity, could trigger multiple outbreak waves unless they are detected timely and accurately (Tao et al. 2021, Oude Munnink et al. 2021).

Although quantitative reverse transcription PCR (RT-qPCR) performs as a gold standard for the SARS-CoV-2 detection, it is barely used to genotype the SARS-CoV-2 VOCs due to the limitations as an amplicon-based method. The current method is heavily based on in-depth whole-genome sequencing, which not only relies on bulk instruments and trained personnel but is also time-consuming and with relatively high cost (Crits-Christoph et al. 2021, Kumar et al. 2021). A recent study showed that the multiplex qPCR could discriminate some SARS-CoV-2 VOCs by targeting specific small deletions(Vogels et al. 2021). However, this method is insensitive to the key point mutations and other developing variations in SARS-CoV-2. Similarly, antigen-based detection assays have lower sensitivity than RT-qPCR and have not yet been reported to identify SARS-CoV-2 VOCs (Peto and Team 2021).

CRISPR-based technology was proven to have unique advantages in molecular diagnosis over the conventional PCR methods in terms of specificity and operability (Chen et al. 2018, Joung et al. 2020, Fozouni et al. 2021). Two CRISPR-based COVID-19 diagnostic methods, DETECTR and SHERLOCK, have been developed using either Cas12a or Cas13a, with the characteristics of rapid, ultrasensitive and easy-to-implement (Broughton et al. 2020, Patchsung et al. 2020). Moreover, CRISPR assays such as RAY were employed to identify the point mutations in SARS-CoV-2 due to the high specificity of Cas/guide-RNA recognition (Kumar et al. 2021, Wang et al. 2021). Nevertheless, the relying on well-designed ligation probes somehow limited the targeting capability to diverse mutation sites. Recently, CRISPR-based point-of-care tests (POCT) of SARS-CoV-2 were reported (de Puig et al. 2021, Teng et al. 2019). The methods provided visual readout using minimal fluorescent instruments and could identify multiple SARS-CoV-2 VOCs, among the synthetic samples though. Hence, significant efforts are still needed in respect to improving the detection sensitivity and validating the methods with clinical samples.

Here, we report a method combining reverse transcription-recombinase polymerase amplification (RT-RPA) and CRISPR-Cas12a technology based on our HOLMES test platform (Li, Cheng, Liu, et al. 2018, Li, Cheng, Wang, et al. 2018), aiming to detect and discriminate SARS-CoV-2 VOCs. By exchanging the crRNA sequences, the method (RRCd) achieved to discriminate the well-known single nucleotide polymorphisms (SNPs) of N501Y and T478K as well as the small deletion of ΔH69-V70, corresponding to the major VOCs worldwide. RRCd was validated in 204 clinical samples after being systematic optimization, showing 99% positive predictive agreement and 100% negative predictive agreement, respectively. The results of RRCd could be visualized within about one hour. Moreover, all procedures including the sample lysis were able to carry out at 37 °C and with no needs for RNA extraction and purification, which is very conducive to the development of POCT products.

## Results

### Technical route of RRCd assay

The CRISPR-*Lb*Cas12a-based nucleic acid test platform HOLMES (Li, Cheng, Wang, et al. 2018) was chosen as the starting core to design the technical route for RRCd (**Figure 1, Supplementary Figure 2**). We conduct a two-step RT-RPA amplification on SARS-CoV-2 genes to provide adequate double-stranded DNA (dsDNA) for the following CRISPR recognition system (**Figure 1, II**). By designing specific crRNA, Cas12a recognizes and *cis*-cleaves the dsDNA templates of SARS-CoV-2 or its variants. Meanwhile, the formation of the ternary complex of Cas12a-crRNA-dsDNA activates the *trans*-activity of Cas12a, which leads to the cleavage of the non-targeted single-stranded DNA (ssDNA) probe in the system (**Figure 1, III**). The results could be quantified directly by fluorescence detection of the fluorophore-quencher ssDNA (FQ-ssDNA) (**Figure 1, IIIa**) or captured by lateral-flow strip using the FITC-Biotin ssDNA (FB-ssDNA) (**Figure 1, IIIb**).

**Figure 1.**
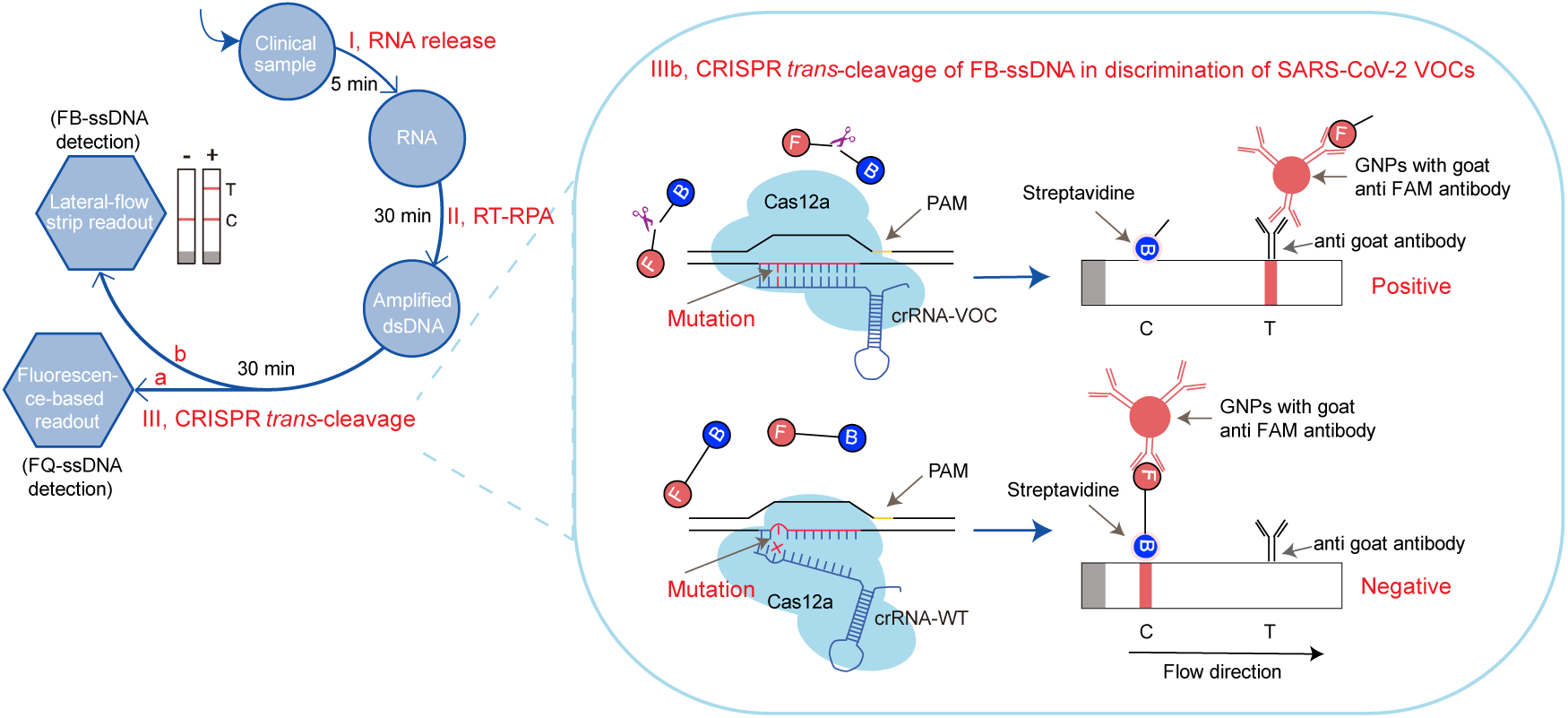
Detection and discrimination of SARS-CoV-2 VOCs by RRCd. Schematic overview of RRCd workflow. (I) SARS-CoV-2 samples were released by automatic extraction (∼10 min) or using RNA release chemical agents without extraction (∼5 min). (II) The viral RNA was reverse transcribed into the dsDNA and amplified by RPA. (III) Cas12a was guided to target the dsDNA by the specific crRNA, thereby activated its *trans*-cleavage activity to the ssDNA probes in the system. The results could be quantified by fluorescence detection of the cleaved FQ-ssDNA (III a), or visualized by lateral-flow strip using the FB-ssDNA probe (III b). RRCd in discrimination of SARS-CoV-2 VOCs with lateral-flow readout is highlighted on the right panel. The target gene from SARS-CoV-2 variant was shown as the template, and it was detected by either crRNA-VOC or crRNA-WT, leading to the positive (upper right panel) or negative (lower right panel) test result, respectively. dsDNA, double-strand DNA; ssDNA, single-strand DNA; FQ, fluorophore (FAM)-quencher (BHQ1); FB, FITC-Biotin; PAM, protospacer adjacent motif; VOC, variant of concern; WT, wild type; GNP, gold nanoparticle; gt, goat.

Hence, different from the previous methods (Broughton et al. 2020, Patchsung et al. 2020, Ding et al. 2020), RRCd combines the advantages of high sensitivity of RT-RPA (Lobato and O’Sullivan 2018) and high specificity of CRISPR-Cas12a (Chen et al. 2018) (**Table 1**). In comparison with Cas9 and Cas13, Cas12a in RRCd possesses greater specificity and lower mismatch tolerance (Chen et al. 2018), making it more suitable for SARS-CoV-2 VOCs genotyping.

**Table 1.**
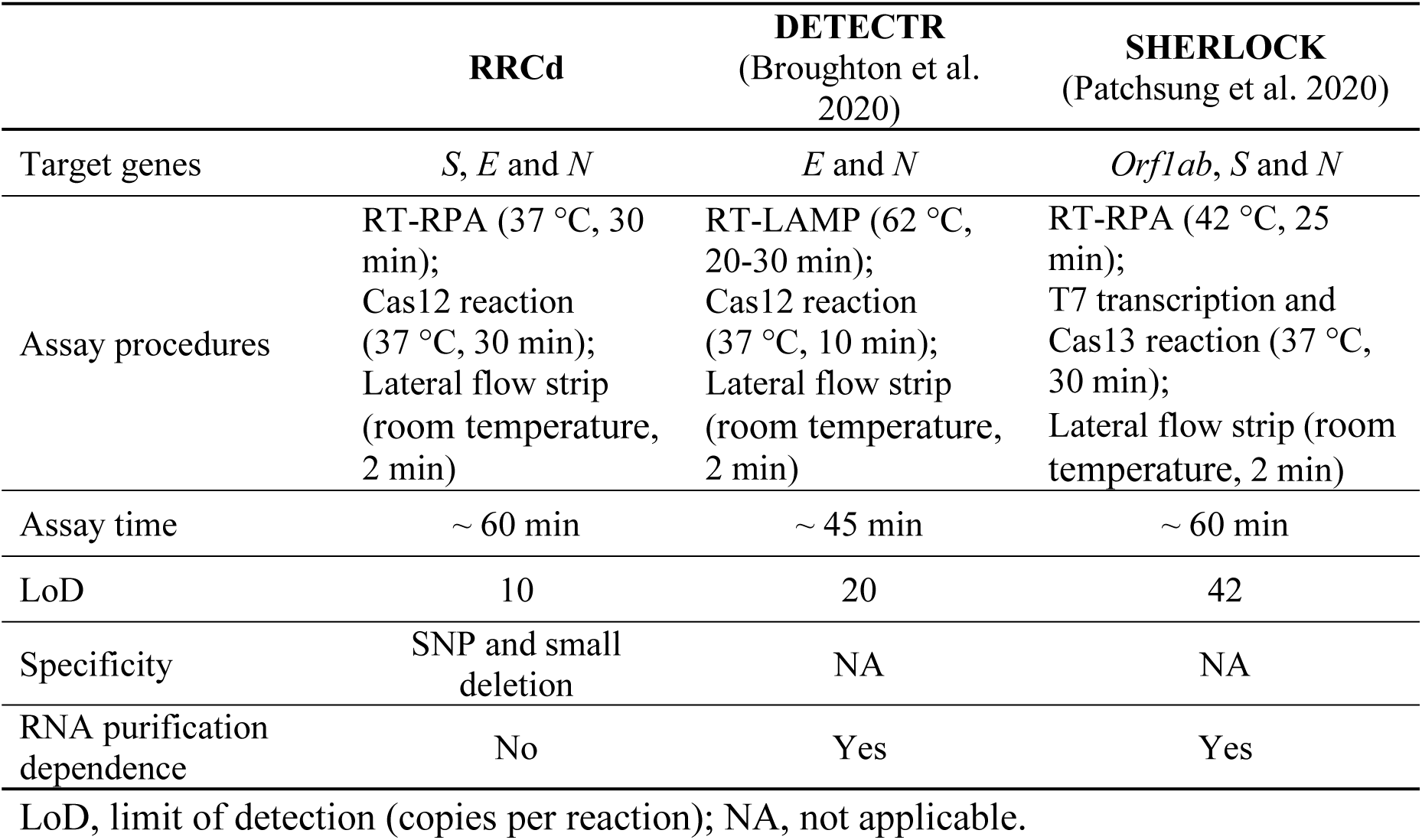
Comparison of RRCd with DETECTR and SHERLOCK for the SARS-CoV-2 detection.

### The specificity analyses of RRCd assay

Representative mutations in spike (S) protein were used for genotyping of the SARS-CoV-2 lineages (Munnink et al. 2021) (**Figure 2A**). By screening optimal RT-RPA primers (**Supplementary Figure 3**) and crRNAs (**Supplementary Figure 4**) that target to the S protein encoding gene, RRCd was readily applied for the detection and discrimination of SARS-CoV-2 VOCs (**Figure 2B-G**). These VOCs include the first reported Alpha and Beta mutation N501Y (Tegally et al. 2021) (**Figure 2B, C**), the recent reported T478K mutation in Delta and Omicron VOCs (Kumar et al. 2022) (**Figure 2D, E**), and the small deletions such as ΔH69-V70 in Alpha and Omicron VOCs (Meng et al. 2021) (**Figure 2F, G**). The N501Y mutation in the receptor-binding domain (RBD) of S protein was reported to cause an increased transmission and infectivity of SARS-CoV-2 (Liu et al. 2022). The T478K nutation in the RBD is also involved in higher transmissibility by the increased affinity to human ACE2 receptor (Kumar et al. 2022), while the deletion of H69V70 at the N-terminal domain is shown to allow the virus to replicate more efficiently (Meng et al. 2021).

**Figure 2.**
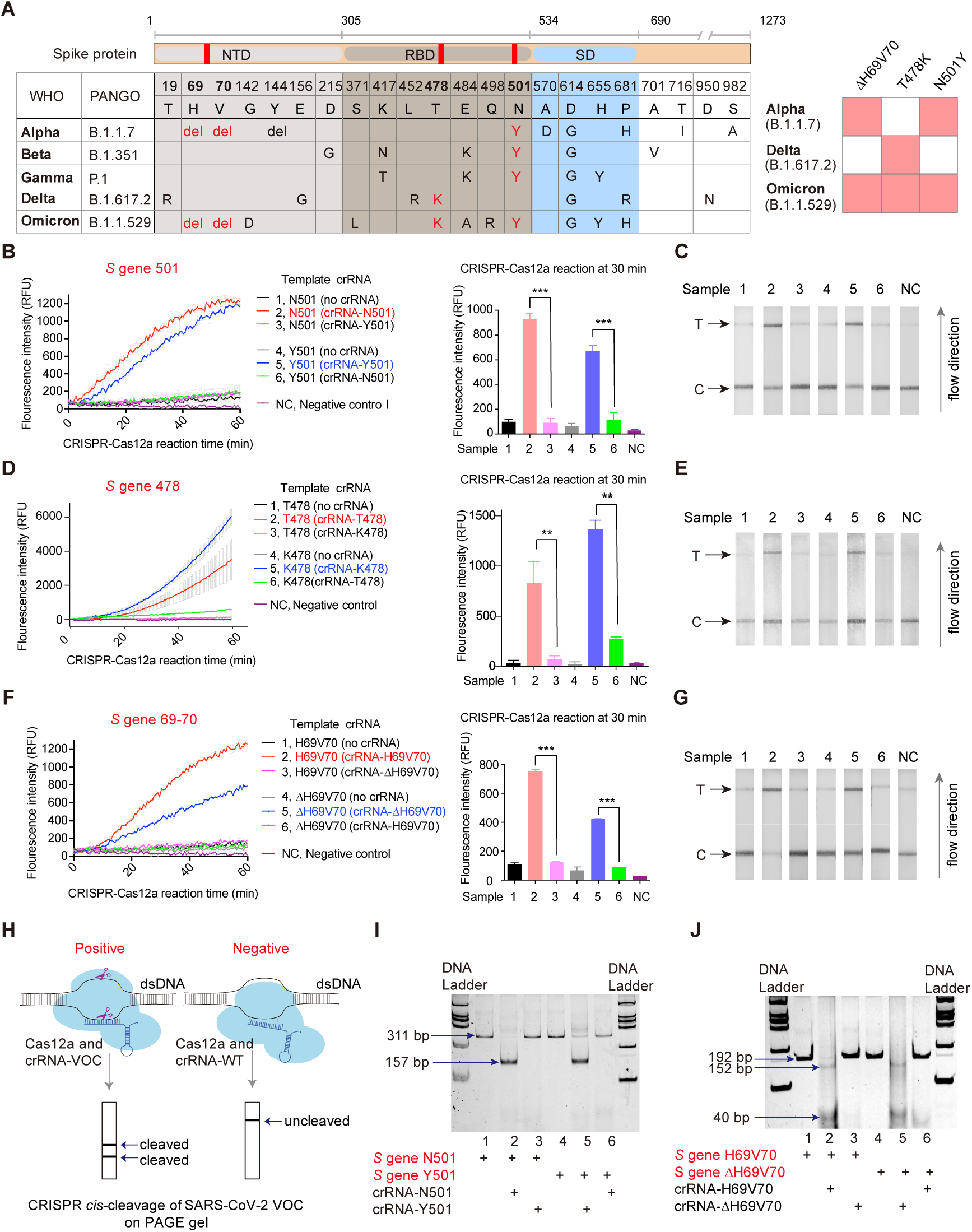
The specificity of RRCd in detection of SARS-CoV-2 VOCs. **A**. Representative changes of amino acids in the spike protein in different SARS-CoV-2 VOCs. Data came from Outbreak.info (https://outbreak.info/compare-lineages) (Julia L. Mullen 2021). The variations detected in this study are colored in red. A simple model is shown on the right panel. Del, deletion. **B**. Specificity analysis of RRCd in detection of SARS-CoV-2 *S* gene 501, using fluorescence-based readouts. The dsDNA derived from pseudovirus *S* gene N501 or Y501 was incubated with Cas12a, FQ-ssDNA, and distinct crRNA at 37 °C, and the corresponding fluorescence signal was recorded over 60 min (left panel) and quantified at 30 min (right panel). Data are presented as the mean ± S.D. (n = 3). ***, *P* < 0.01 by student’s *t* test. RNase-free water was used as the negative control (NC). RFU, relative fluorescence units. **C**. Specificity analysis of RRCd using lateral-flow readouts. The samples analyzed were the same as those in panel B. The reactions were conducted at 37 °C for 30 min before the mixtures were transferred onto the strips, using FB-ssDNA probe instead of FQ-ssDNA. The relative greyness (G) of strip bands was quantified using ImageJ. In this study, G_Sample_ / G_NC_ of the T band ≥ 1.5 indicates positive. **D, E**. Specificity analysis of RRCd in detection of SARS-CoV-2 *S* gene 478. **F, G**. Specificity analysis of RRCd in detection of SARS-CoV-2 *S* gene 69-70. **H**. Schematic of RRCd in detection of SARS-CoV-2 VOCs by Cas12a *cis*-cleavage activity. **I, J**. The specificity was demonstrated by PAGE analyses, using SARS-CoV-2 variants of N501Y and ΔH69-V70 as the examples.

Our test results were demonstrated by both the fluorescence-based and lateral-flow readouts, representing the *trans*-cleavage activity of Cas12a (Chen et al. 2018, Li, Cheng, Liu, et al. 2018) (**Figure 2B-G**). Additionally, we confirmed this specific cleavage using polyacrylamide gel electrophoresis analyses by leveraging of the Cas12a *cis*-cleavage activity (Chen et al. 2018) (**Figure 2H-J**). Moreover, RRCd can be used for the discrimination of SARS-CoV-2 from other phylogenetic related viruses (**Supplementary Figure 5**).

### The detection limit of RRCd

To determine the limit of detection (LoD) of RRCd, we built a standard curve to dissect the correspondences between the viral copy numbers and the *C*_t_ values using SARS-CoV-2 pseudovirus (**Supplementary Figure 6**). The *C*_t_ values of each dilution were measured using RT-qPCR targeting *N* gene or *ORF1ab* gene to mimic the clinical *C*_t_ ranges. Despite showing distinct kinetics, the fluorescence-based readouts revealed that the LoDs of all the targeted regions (*E* gene, N501, Y501, T478, and K478 of *S* gene) were ∼10 copies per reactions, corresponding to a *C*_t_ value of 35 in the RT-qPCR assay (**Figure 3**). The fluorescence results from a 30-min CRISPR reaction suggested that the detection sensitivity of N501 is a slightly higher than that of Y501 (**Figure 3B, C**), while the detection sensitivity of T478 is a slightly lower than that of K478 (**Figure 3D, E**). It is worth noting that the detection sensitivity of the lateral-flow readouts is about 10-folds lower than that of the fluorescence-based readouts (**Figure 3**). The detection of Y501 and K478 was shown to be the most sensitive when using the lateral-flow readout, with the LoDs of ∼10 copies per reactions (**Figure 3C, E**).

**Figure 3.**
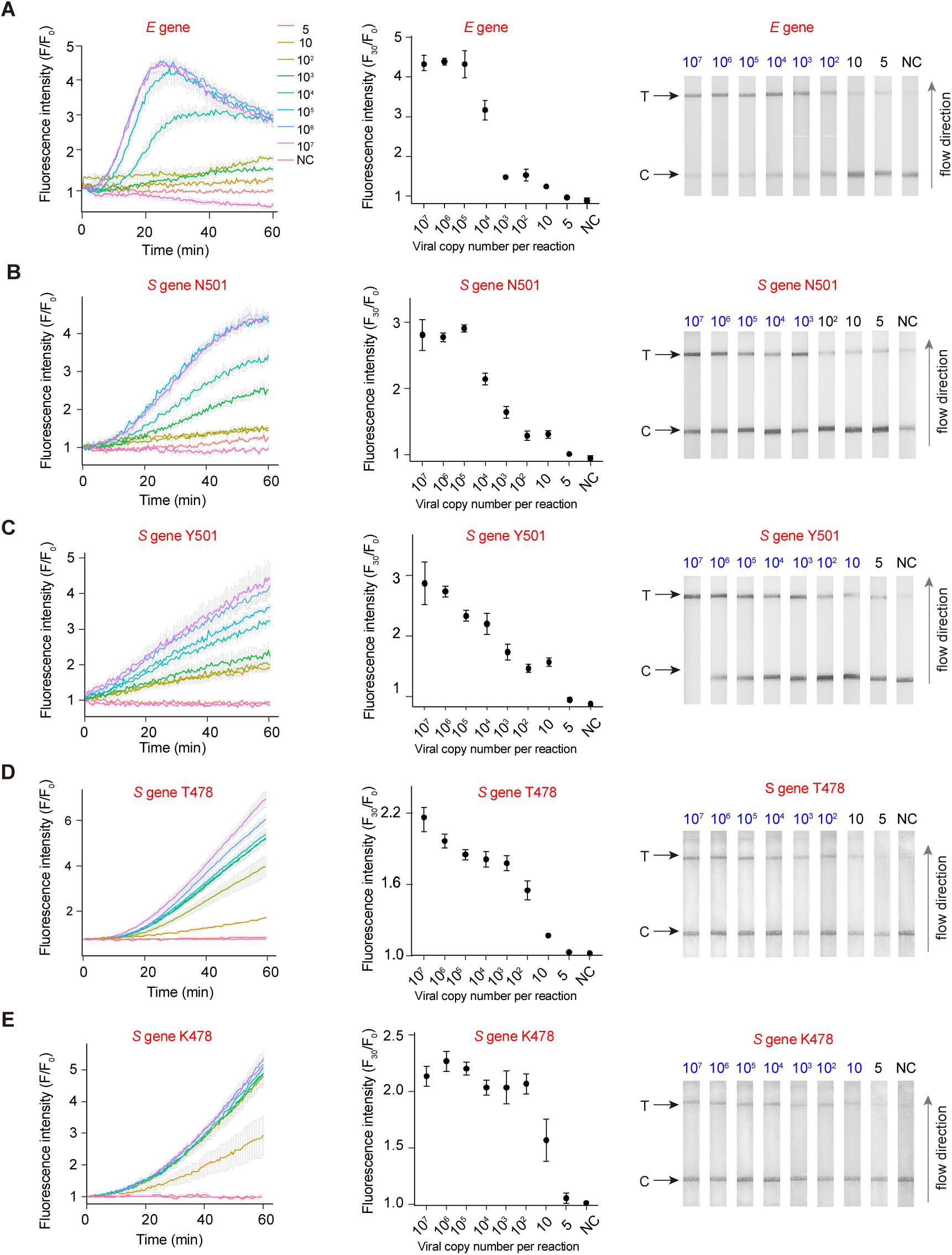
The detection limit of RRCd for SARS-CoV-2 VOCs. The detection limit of the targeted regions in VOCs were determined by using 10^7^, 10^6^, 10^5^, 10^4^, 10^3^, 10^2^, 10 and 5 copies of pseudovirus RNA per reaction, respectively. The dsDNA derived from the pseudovirus RNA was incubated with Cas12a, ssDNA probe, and crRNA at 37 °C for 60 min (fluorescence-based readouts) or 30 min (lateral-flow readouts), respectively. The relative fluorescence intensity (F/F^0^) was calculated as the fluorescence signal versus the starting signal (left panel). Correspondingly, the fluorescence signal at the 30 min-reaction was quantified (middle panel). The positive samples of lateral-flow readouts (right panel) are indicated as blue by the T-line quantification as described. Data are presented as mean ± S.D. from at least three independent experiments. For the dilutions with 10 and 5 copies per reaction, at least five independent experiments were conducted. **A**. LoD determination of RRCd for *E* gene. **B-E**. LoD determinations of RRCd for N501 (B), Y501 (C), T478 (D) and K478 (E) in *S* gene, respectively. NC, negative control using the RNase-free water.

To avoid false-discriminative results, we further assessed the factors that may affect the detection sensitivity in RRCd, including the sample volume and probe inputs, the usage of RNase H, T4 gene 32 protein, Bsu DNA polymerase, RPA kits, and Mg^2+^. We found the sample volume and the usage of T4 gene 32 protein (Villalva et al. 2001) are the key factors affecting RRCd sensitivity mostly (**Supplementary Figure 7**).

### Validation of RRCd in detection and discrimination of clinical samples

Based on the optimized procedures, we evaluated the performance of RRCd on detecting COVID-19 clinical samples isolated from nasopharyngeal and salivary swabs. In coordination with the scenario of POCT, our available samples were collected from Shanghai Customs where the susceptible patients were characterized as asymptomatic and with low virus load (**Supplementary Figure 8**). To minimize the cognitive bias, all information of the clinical samples was confidential to study staff before the detection.

A validation study of SARS-CoV-2 detection was conducted on a total of 96 clinical samples (54 RT-qPCR-verified positive samples and 42 RT-qPCR-verified negative samples). Both *E* gene and *S* gene were used as the targeted sequences to acquire the double-checked detection results (**Figure 4, Supplementary Figure 9**). RRCd identified all the positive samples with *C*_t_ < 33 through both lateral-flow and fluorescence-based readouts. In addition, 42 COVID-19 negative samples were all identified without false positive results from both readouts. Samples beyond the *C*_t_ value of 32 were more likely to be detected by targeting the *E* gene (*C*_t_ ≤ 35, 100% detected) than the *S* gene (**Figure 4**). The detection sensitivity of these results was comparable to the RT-LAMP-Cas12 based DETECTR (Broughton et al. 2020) and the RT-RPA-Cas13 based SHERLOCK (Patchsung et al. 2020), showing a 99% positive predictive agreement (PPA) and 100% negative predictive agreement (NPA) in both readouts (*C*_t_ < 33) (**Table 2**).

**Figure 4.**
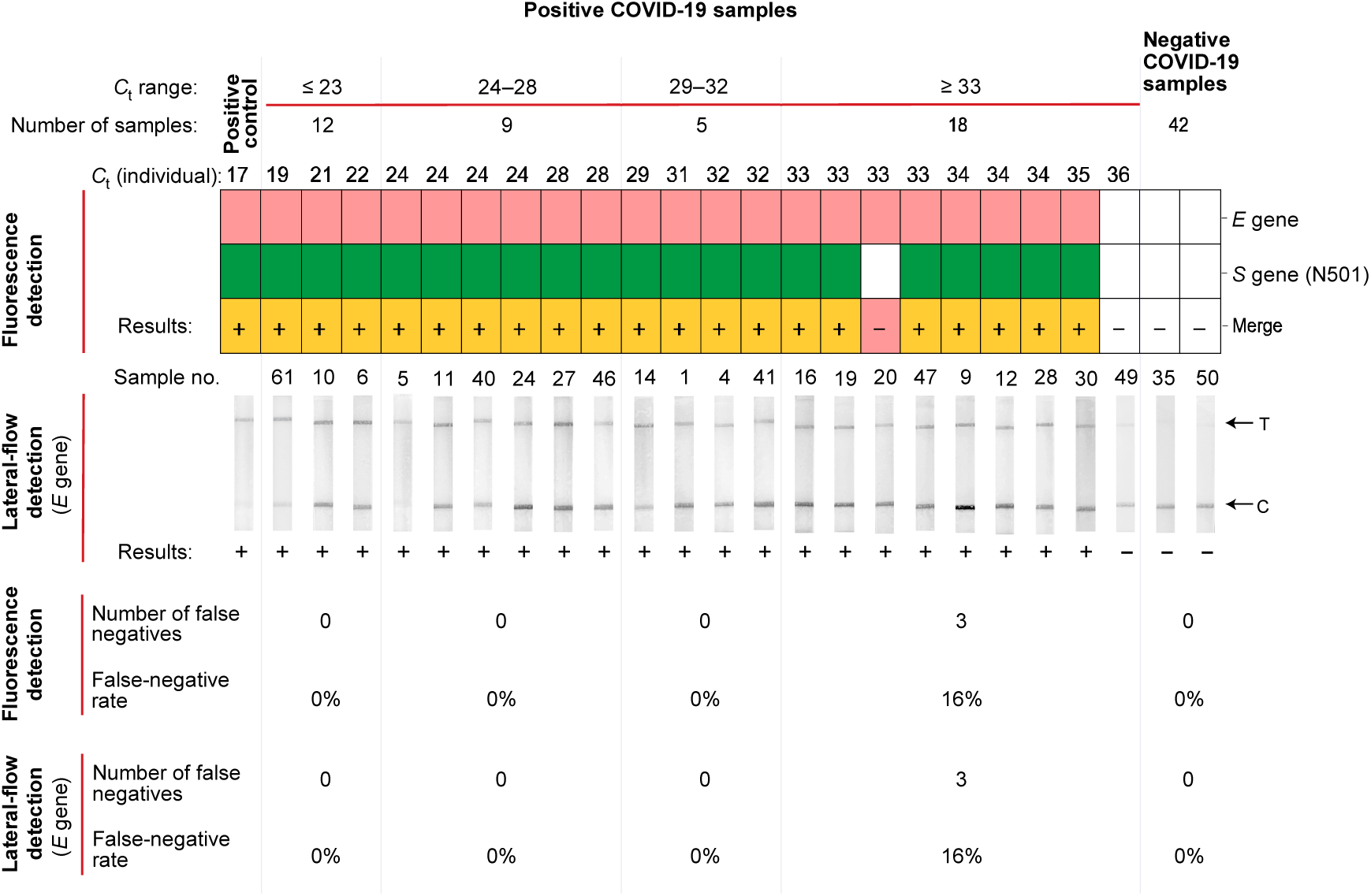
Validation of RRCd in detection of 96 RT-qPCR-verified clinical samples. Nasopharyngeal and salivary swabs were collected from 20^th^ January to 10^th^ April 2021 at Shanghai Customs, and a double-checked detection (*E* gene and *S* gene) was executed for them by RRCd. To minimize the cognitive bias, all information of the clinical samples was confidential to study staff before the detection. The results are displayed according to the *C*_t_ values of COVID-19 positive samples, and the total number of clinical samples in each *C*_t_ range are shown on the top of the panel. Representatives of fluorescence-based readouts and lateral-flow readouts of the positive samples as well as the negative samples are shown on the middle of the panel. Both *E* gene and *S* gene (N510) were used as the targeted sequences in fluorescence-based readouts. The light red and green indicate the positive detection results of *E* gene and *S* gene (N501), respectively, while the orange represents the double-checked positive result. The white indicates the negative result. Only the lateral-flow results of *E* gene are shown. +, positive;–, negative or indistinguishable. The analyses of the detection results are shown as the number of false negatives and the false-negative rate for each *C*_t_ range on the bottom of the panel. The SARS-CoV-2 pseudovirus was used as the positive control. The full dataset is shown in Supplementary Figure 9.

**Table 2.** Predictive agreements of RRCd in detection of clinical samples.

|  | RRCd results | RT-qPCR-verified positive samples |  | RT-qPCR-verified negative samples |
| --- | --- | --- | --- | --- |
| | | $C_t < 33$ | $C_t \geq 33$ | |
| Fluorescence-based readout | Positive | 84 (true positive) | 24 (true positive) | 0 (false positive) |
|  | Negative | 1 (false negative) | 13 (false negative) | 82 (true negative) |
|  | Total | 85 | 37 | 82 |
|  | PPA/NPA | 99% | 65% | 100% |
|  | Total PPA/NPA | 89% |  | 100% |
| Lateral-flow readout | Positive | 84 (true positive) | 23 (true positive) | 0 (false positive) |
|  | Negative | 1 (false negative) | 14 (false negative) | 82 (true negative) |
|  | Total | 85 | 37 | 82 |
|  | PPA/NPA | 99% | 62% | 100% |
|  | Total PPA/NPA | 88% |  | 100% |

In total, 204 clinical samples including 122 positive samples and 82 negative samples were analyzed by RRCd with both fluorescence-based and lateral-flow readouts. PPA, positive predictive agreement; NPA, negative predictive agreement.

We next evaluated the performance of RRCd on discrimination of SARS-CoV-2 VOCs. The 501 position of *S* gene was used as a target to detect SARS-CoV-2 VOCs, such as the Alpha, Beta, Gamma, and Omicron (Tegally et al. 2021). A total of 68 clinical samples, consisting of 28 genome-sequencing-verified N501 samples and 19 genome-sequencing-verified Y501 samples, were collected from March 3^rd^ to April 10^th^ 2021 at Shanghai Customs. RRCd discriminated all the variants from samples with a *C*_t_ < 32, either using lateral-flow readouts or fluorescence-based readouts (**Figure 5A, Supplementary Figure 10**). However, the discrimination ability decreased strikingly when testing the samples with *C*_t_ values beyond 33, which is possibly due to the relatively high LoD of *S* gene 501 as mentioned above (**Figure 3**).

**Figure 5.**
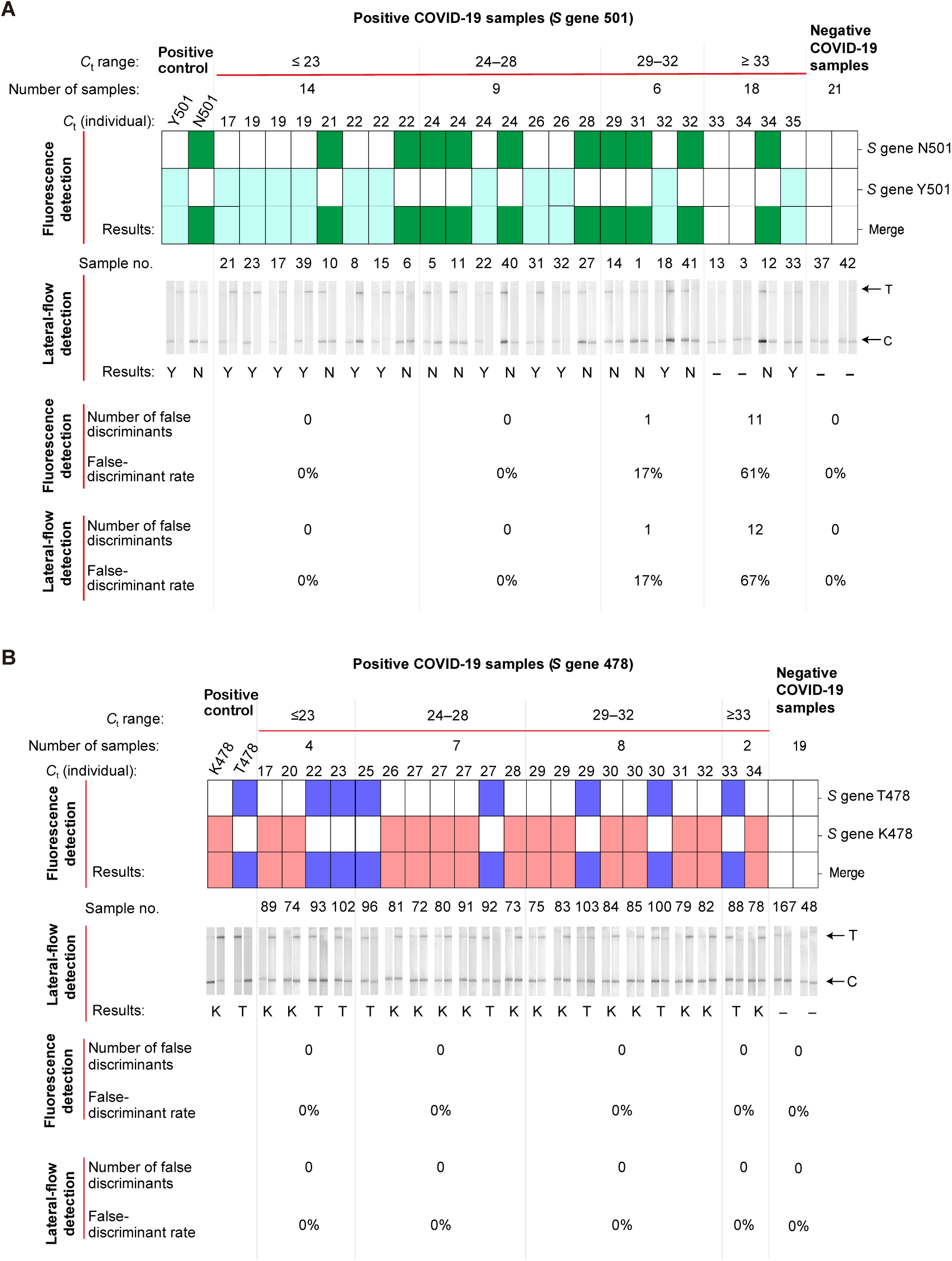
Validation of RRCd in detection and discrimination of 108 genome-sequencing-verified clinical samples. **A**. Discrimination of SARS-CoV-2 *S* gene 501 variants in clinical samples. The total number of clinical samples in each *C*_t_ range are shown on the top of the panel. In total, 47 positive samples (*C*_t_ range of 13-35) and 21 negative samples were detected. The *S* gene 501 variants were discriminated by crRNA-N501 (green) and crRNA-Y501 (light blue) using fluorescence-based readouts. In addition, the lateral-flow readouts are displayed in the middle of the panel using crRNA-N501 (left) and crRNA-Y501 (right). N, N501; Y, Y501; –, negative or indistinguishable. The analysis results are shown as the number of false discriminants and the false-discriminant rate for each *C*_t_ range on the bottom of the panel. **B**. Discrimination of SARS-CoV-2 *S* gene 478 variants in clinical samples. In total, 21 positive samples (*C*_t_ range of 17-34) and 19 negative samples were detected. The *S* gene 478 variants were discriminated by crRNA-T478 (blue) and crRNA-K478 (pink) by fluorescence readouts. The lateral-flow readouts using crRNA_T478 (left) and crRNA_K478 (right) are displayed on the below. T, T478; K, K478;–, negative or indistinguishable. The corresponding SARS-CoV-2 pseudoviruses were used as the positive controls. The full datasets for panel a and b are shown in Supplementary Figure 10 and Supplementary Figure 11, respectively.

The 478 position of *S* gene could be used as a target to identify Delta and Omicron VOCs (**Figure 5B, Supplementary Figure 11**). A total of 40 clinical positive samples, consisting of 7 genome-sequencing-verified T478 samples and 14 genome-sequencing-verified K478 samples, were collected from May 27^th^ and October 29^th^ 2021. RRCd was able to discriminate all the variants targeting the *S* gene 478. Moreover, RRCd displayed a relatively higher sensitivity in detecting the position of 478 than that of 501, with the *C*_t_ value reaching 34. Although using a relatively small sample group, the detection results were confirmed by both readouts with 0% false-discriminant rates (**Figure 5B**). Taken together, these clinical data suggested that RRCd was able to detect and discriminate the SARS-CoV-2 VOCs, accurately and sensitively (**Table 2**).

## Discussion

Although a decreased lethality of the circulating Omicron has been reported, its overall mortality rate has not dropped because its dramatically increased infectivity (Nyberg et al. 2022). The frequent outbreak waves of COVID-19 highlight the needs for detection methods that are rapid, sensitive, and specific. However, the current method relying on deep genome sequencing is limited by the high cost, long turn-around time, and the reduced overall sensitivity (Kumar et al. 2021). Although the recently developed CRISPR-based methods were characterized as sensitive and easy-to-implement, most of them were barely validated by the large clinical samples or have a limited adaptability (Wang et al. 2021, Teng et al. 2019, de Puig et al. 2021).

In the present study, RRCd completed in ∼1 hour and displayed a sensitivity down to 10 copies per reaction. By targeting and identifying the representative N501Y, T478K and ΔH69-V70 mutations, RRCd discriminated COVID-19 samples of Alpha, Delta, and Omicron VOCs. The method was validated in 204 clinical COVID-19 samples, with 99% PPA and 100% NPA for the samples of *C*_t_ < 33 through both the fluorescence and the lateral-flow readouts (**Table 2**). Moreover, The SARS-CoV-2 detection by direct lysing the clinic samples without RNA extraction and purification was demonstrated (**Supplementary Figure 12**). Although a relatively small sample size was used, these results indicated that the whole process of RRCd including the RT-RPA amplification and the lateral-flow detection is independent of electrical devices, which may facilitate the development of POCT products (**Table 1**).

In addition, the RRCd assay has the potential to be easily adapted to detect novel mutations on SARS-CoV-2 or other viruses by designing the new RT-RPA primers and crRNAs, without changing the subsequent detection procedures and ssDNA probes (**Figure 1, Supplementary Figure 2**). Overall, RRCd can be performed in sites with few detection equipment (*e*.*g*., self-testing) or limited power access (*e*.*g*., underdeveloped areas), or situations when quick turnaround times are needed (*e*.*g*., hospital emergency). Nevertheless, the POCT products for RRCd needs to be further investigated.

## Materials and methods

### Clinical sample collection and ethics statement

All 204 nasopharyngeal and salivary samples were collected at Shanghai International Travel Healthcare Center (SITHC), Shanghai Customs District P. R. China, from March 3^rd^ and to October 29^th^ 2021. The samples were treated in strict accordance with WHO recommended standard operation procedure. Ethical approval of the study was given by the Bioethics Committee of Bio-X Institute of Shanghai Jiao Tong University (COA: M202007).

### RT-qPCR for the SARS-CoV-2 detection

The viral RNA of each swab was extracted and purified from a 200 μl-swab transport media using an automatic nucleic acid platform (SSNP-3000A, Bioperfectus tech., China) according to the manufacturer’s instruction. A 100 μl RNase-free water was used to elute the RNA. Viral lysis and RNA extraction were performed in a biosafety level 3 laboratory, whereas other procedures were performed in a biosafety level 2 laboratory.

The quantitative reverse transcription PCR (RT-qPCR) for SARS-CoV-2 detection was performed with a 2019-nCoV nucleic acid detection kit (BioGerm, Shanghai, China) according to the manufacturer’s protocol. Each RT-qPCR reaction had a final volume of 25 μl, including 5 μl purified RNA, 12 μl reaction buffer, 4 μl enzyme solution and 4 μl probe primers solution. RT-qPCR assay was performed on QuantStudio Dx Real-Time PCR (Applied Biosystems, MA, USA) under the following conditions: incubation at 50 °C for 15 min and 95 °C for 5 min, 45 cycles of denaturation at 94 °C for 15 s, and extension and fluorescence signal reading at 55 °C for 45 s. Two target genes, *i*.*e*., the *ORF1ab* gene and the *N* gene, were simultaneously amplified and tested in the RT-qPCR assay. The test result was deemed as positive when the cycle threshold values (*C*_t_ value) of both *ORF1ab* and *N* genes ≤ 38; otherwise, a negative result was deemed. These diagnostic criteria were based on the recommendation from the National Institute for Viral Disease Control and Prevention (China) (http://ivdc.chinacdc.cn/kyjz/202001/t20200121_211337.html).

### Design and screening of the RT-RPA primers and crRNAs

For the detection of SARS-CoV-2, *E* gene was selected as the test target using the RT-RPA primers and crRNA recommended by WHO with minor modifications (**Supplementary Table 1**).

For the detection of SARS-CoV-2 VOCs, the representative mutations in the *S* gene were selected as the test targets. Aiming to distinguish the most prevailing SARS-CoV-2 VOCs (*i*.*e*., the Alpha, Delta, and Omicron), we designed the RT-RPA primers on the conserved regions flanking the mutations of *S* gene 478 and 501 (**Supplementary Table 1**), respectively, and screened the best pairs by agarose gel analyses for the amplification efficiency of dsDNA. Two pairs of optimal RT-RPA primers (S501RPAF1/S501RPAR1 and S478RPAF3/S478RPAR5) were selected for the detection of *S* gene 478 and 501, respectively. To further evaluate their applicability in the detection of clinical samples, these RT-RPA primers were aligned with 859,132 full-length SARS-CoV-2 genome sequences collected from January 1^st^ 2021 to December 31^th^ 2021 in the GISAID database (Buch et al. 2011). The results showed that these pairs of RT-RPA primers hit 99.7% of the genome sequences, demonstrating a good applicability in detection of prevailing clinical samples.

Optimal crRNAs for the discrimination of *S* gene 501 variants were assessed by determining the specificity of the crRNA to the single-base mismatched *S* gene templates. DNA fragments containing wild-type *S* gene 501 with point mutations were synthesized in pUC19 backbone plasmid by Sangon Biotech. These *S* gene 501 regions were transcribed into RNA using HiScribe T7 High Yield RNA Synthesis kit (Cat^#^ E2050S, New England Biolabs) at 37 °C for 4 h, whose products were further analyzed using both fluorescence-based and lateral-flow readouts. The crRNA-N501 was employed to discriminate the single-base mismatches. Based on this approach, we designed the crRNAs for the discrimination of other SARS-CoV-2 VOCs. All crRNA and RT-RPA primer sequences used in this study are listed in **Supplementary Table 1**.

### crRNA generation

The DNA templates (**Supplementary Table 1**) for *in vitro* transcription (IVT) of crRNAs were designed with T7 primer binding sites using CRISPR-RT Design Tool (http://bioinfolab.miamioh.edu/CRISPR-RT) and synthesized by Sangon Biotech (Shanghai, China). The crRNAs were transcribed using HiScribe T7 High Yield RNA Synthesis kit (Cat^#^ E2050S, New England Biolabs) at 37 °C for 16 h.

Then, the crRNAs were purified using RNA Clean and Concentrator kit (Cat^#^ R1017, Zymo Research). The concentration of crRNA was quantified by NanoDrop 2000C spectrophotometer (Thermo Scientific) and stored at -80 °C before use.

### Reverse transcription and recombinase polymerase amplification (RT-RPA)

A 20 μl volume-basic RT reaction mixture was used with 1 × RT buffer (Cat^#^ EP0442, Thermo Scientific), 10 μl RNA template, 200 U RevertAid reverse transcriptase (Cat^#^ EP0442, Thermo Scientific), 1.25 μM RT-RPA forward and reverse primer mix, 1 mM dNTP mix (Cat^#^ P031-01, Vazyme, Nanjing, China), and 20 U RNase inhibitor (Cat^#^ N8080119, Thermo Scientific). The RT reaction mixture was incubated at 37 °C for 15 min to generate cDNA. Then, a 10 μl RT product was transferred into the 50 μl-basic RPA reaction mixture, which contains 29.5 μl rehydration buffer, one freeze-dried RPA reaction pellet (Cat^#^ TABAS03KIT, TwistDx), 0.5 μM RT-RPA forward and reverse primer mix, and 14 mM MgOAc (last added to initiate the reaction). The RPA reaction mixture was incubated at 37 °C for another 15 min. The RNA extracts from SARS-CoV-2 pseudovirus and the RNase-free water were used as the positive control (PC) and negative control (NC), respectively.

### CRISPR *trans*-cleavage assay for fluorescence-based and lateral-flow readouts

Briefly, a 100 μl volume basic CRISPR reaction mixture was used with 1 × *Lba*Cas12a buffer, 5 μl RT-RPA product, 0.2 μM *Lba*Cas12a (Cat^#^ M0653T, New England Biolabs), 0.6 μM crRNA, 10 μM MgCl_2_, 10 nM fluorophore (FAM)-quencher (BHQ1) ssDNA probe (FQ-ssDNA, Sangon Biotech) for the fluorescence-based detection, or 50 nM FTIC-Biotin ssDNA probe (FB-ssDNA, Zoonbio Biotechnology, Nanjing, China) for the lateral flow-based detection.

For the fluorescence-based readout, a 96-well plate containing 100 μl reaction mixture was incubated at 37 °C using the Cytation 5 Mul-Mode Reader (BioTek, VT, USA). Usually, a 60-min monitoring was used with an interval time of 30 s. The fluorescence excitation and emission wavelength were set as 485 nm and 520 nm, respectively. The relative fluorescence intensity (F/F_0_) was calculated as the fluorescence signal relative to the starting signal. Generally, the fluorescence signals at the 30 min-reactions were quantified as F_30_/F_0._ In this study, (F_30_/F_0_)_Sample_ / (F_30_/F_0_)_NC_ ≥ 1.2 indicates the positive result while (F_30_/F_0_)_Sample_ / (F_30_/F_0_)_NC_ < 1.2 indicates the negative result.

For the lateral flow readout, a 100 μl-reaction mixture was incubated in an Eppendorf tube at 37 °C for 30 min. Then, the HybriDetect strip (Milenia Biotec, Giessen, Germany) was placed into the Eppendorf tube, and the C-line and/or T-line were visualized and stabilized in ∼2 min. The relative greyness of the T-line (G) was quantified using ImageJ. In this study, G_Sample_ / G_NC_ ≥ 1.5 indicates the positive result while G_Sample_ / G_NC_ < 1.5 indicates the negative result. Due to the incomplete digestion of FB-ssDNA, some positive test results displayed two lines (T-line and C-line) on the strips.

### CRISPR *cis*-cleavage assay

Upon detection of the targeted sites, the CRISPR-Cas12a system is capable to *cis*-cleave the dsDNA template into two short fragments, the activity of which was determined by sodium dodecyl sulfate-polyacrylamide gel electrophoresis (SDS-PAGE). After a 30-min incubation of the CRISPR reaction mixture as described above, the mixture was denatured at 95 °C for 5 min. Then, it was loaded into an 8% SDS-PAGE for 45 min at 150 V. The gel was stained with GelRed (Biotium, CA, USA) and analyzed by Image J.

### Specificity of RRCd assay

To test if RRCd could discriminate SARS-CoV-2 from the phylogenetically related viruses, DNA fragments containing selected regions of *N* gene of SARS-CoV (NC_004718.3), MERS CoV (NC_019843.3), and H1N1 (NC_026433.1, NC_026434.1) were synthesized from Sangon Biotech (**Supplementary Table 2**), and then cloned into a T7 promoter expression plasmid to be used as the templates for IVT of viral RNAs, respectively. The transcribed RNAs were detected using crRNA-SARS-CoV2 *N* by the RRCd assay as described above.

### Building a standard curve to determine the relationship between viral copy numbers and the *C*_t_ values

The pseudovirus SARS-CoV-2 ab II EMNS S-MT and SARS-CoV-2-MT-B.1.617.2. (**Supplementary Table 3**) were purchased from FUBIO, Nanjing, China. To establish the correlation between viral copy numbers and the *C*_t_ values of SARS-CoV-2, we built a standard curve using a gradient dilution (1 × 10^5^, 1 × 10^4^, 1 × 10^3^, 1 × 10^2^, 10 and 5 RNA copies per reaction) of SARS-CoV-2 ab II EMNS S-MT. The *C*_t_ values of the samples were determined by targeting *N* gene and *ORF1ab* gene with RT-qPCR as mentioned above. Results were plotted against the viral copy numbers yielding a linear relationship.

### Limit of detection (LoD) of RRCd assay

To determinate the detection limits of RRCd assay for different SARS-CoV-2 variants (*E* gene, T478, K478, N501 and Y501 in *S* gene), nine serial dilutions (1 × 10^7^, 1 × 10^6^, 1 × 10^5^, 1 × 10^4^, 1 × 10^3^, 1 × 10^2^, 10 and 5 copies per reaction) of SARS-CoV-2 pseudovirus RNAs were used and their fluorescence-based and lateral-flow readouts were analyzed as described above. For the dilutions (1 × 10^7^, 1 × 10^6^, 1 × 10^5^, 1 × 10^4^, 1 × 10^3^ and 1 × 10^2^ copies per reaction), at least three independent experiments were performed, while for the dilutions with 10 and 5 copies per reaction, at least five independent experiments were conducted. RNase-free water was used as the negative control.

### Whole-genome sequencing of clinical samples

To determine the lineages of the RT-qPCR-verified positive clinical samples, whole-genome sequencing of SARS-CoV-2 cDNA was conducted by Illumina Next-generation sequencing. The first-strand cDNA of SARS-CoV-2 RNA was generated using the SuperScript IV First-Strand Synthesis System (Cat^#^ 18091050, Thermo Fisher Scientific). The corresponding libraries were constructed using the cDNAs with the ARTIC SARS-CoV-2 amplification protocol (as described in https://artic.network/ncov-2019). Paired-end 150 bp sequencing was performed for each library on Illumina X-10. Genome assembly was done using CLC Genomics Workbench (http://www.clcbio.com/products/clcgenomics-workbench/) (Liu and Di 2020). The Phylogenetic Assignment of Named Global Outbreak Lineages (PANGOLIN) (https://virological.org/t/pangolin-web-application-release/482) (Lam et al. 2020) and the NextClade (https://clades.nextstrain.org/) (Hadfield et al. 2018) were used for lineage classification and clade classification, respectively. All the *S* gene from the positive clinical samples used in this study were re-sequenced by Sanger method.

### Optimization of RRCd assay

To optimize the RT reaction, we first investigated the effects of different RNA input (1, 5, or 10 μl) on the detection sensitivity of RRCd. RNA was extracted from a positive clinical sample (*C*_t_ = 30) as described above and the SARS-CoV-2 *E* gene was used as the test target.

RNase H is an endoribonuclease that specifically hydrolyzes the RNA-DNA hybrids(Qian et al. 2020), while T4 gene 32 protein is known to increase the efficiency of reverse transcriptase(Villalva et al. 2001) and Bsu DNA polymerase has the strand displacement DNA synthesis activity(Piepenburg et al. 2006). To evaluate the effects of these enzymes on RT-RPA efficiency, a basic RT reaction mixture was prepared as described above with or without the addition of 250 U RNase H (Cat^#^ M0297S, New England Biolabs), 16.5 μM T4 gene 32 protein (Cat^#^ M0300L, New England Biolabs) or 0.25 U Bsu DNA polymerase (Cat^#^ M0330S, New England Biolabs), respectively.

The corresponding RT products were used as inputs to the RPA assay and subsequent products were analyzed by fluorescence based CRISPR assay as described above. Finally, a 20 μl volume-optimized RT reaction mixture was obtained as below: 1 × RT buffer, 10 μl template RNA, 200 U RevertAid reverse transcriptase (Thermo Scientific), 1.25 μM RT-RPA forward and reverse primer mix, 1 mM dNTP mix, 20 U RNase inhibitor (Thermo Scientific), 250 U RNase H (New England Biolabs) and 16.5 μM T4 gene 32 protein (New England Biolabs).

For screening of an optimal RPA kit, four RPA assay kits were compared under the same conditions, including Amp-Qitian kit (Qitian gene Biotech, Wuxi, China), Amp-Future kit (Amp-Future, Weifang, China), TwistAmp Baic kit (TwistDx, Cambridge, UK), and Amp-Yizhi Kit (Yizhi Biotech, Shenzhen, China). Briefly, 10 μl RT product was transferred into a 50 μl volume basic RPA reaction mixture using these four kits, respectively, which contains 29.5 μl rehydration buffer, one freeze-dried RPA reaction pellet, 0.5 μM RT-RPA forward and reverse primer mix, and 14 mM MgOAc (last added to initiate the reaction). The RPA reaction mixture was incubated at 37 °C for 15 min. RNase-free water was used as the negative control.

To investigate the possible effects of T4 gene 32 protein and Bsu DNA polymerase on RPA reaction, basic RPA reaction mixture was prepared as described above with or without the addition of 16.5 μM T4 gene 32 protein (Cat^#^ M0300L, New England Biolabs) or 0.25U Bsu DNA polymerase (Cat^#^ M0330S, New England Biolabs).

The RT-RPA products were analyzed by fluorescence based CRISPR assay as described above. Finally, a 50 μl volume-optimized RPA reaction mixture was obtained as below: 29.5 μl rehydration buffer (TwistDx), one freeze-dried RPA reaction pellet (TwistDx), 10 μl RT product, 0.5 μM RT-RPA forward and reverse primer mix, 16.5 μM T4 gene 32 protein (Cat^#^ M0300L, New England Biolabs) and 14 mM MgOAc.

To determine if the CRISPR reaction has a higher efficiency using manganese rather than magnesium, as described before(Li et al. 2020). The fluorescence-based CRISPR assay was used to quantify the results as described above, with 10 mM MnCl_2_ or MgCl_2_ in the basic CRISPR reaction mixture.

Furthermore, the optimal concentrations of FQ-ssDNA and FB-ssDNA probes were determined within the basic CRISPR reaction mixture. The FQ-ssDNA probe (10, 15, or 20 nM) and FB-ssDNA probe (0.5, 5, 25, 50, 125, 200, 500, 1000, or 2000 nM) were used in the CRISPR reaction, and the corresponding reaction fluorescence intensities and lateral-flow readouts were recorded as described above, respectively.

Finally, a 100 μl volume-optimized CRISPR reaction mixture was obtained as below: 1 × *Lba*Cas12a buffer (Cat^#^ M0653T, New England Biolabs), 5 μl RT-RPA product, 0.2 μM *Lba*Cas12a (Cat^#^ M0653T, New England Biolabs), 0.6 μM crRNA, 10 μM MgCl_2_, and 20 nM FAM-BHQ1 ssDNA probe for the fluorescence-based detection, or 200 nM FTIC-Biotin ssDNA probe for the lateral flow-based detection.

### Comparison of the one-step and two-step RT-RPA

The amplification efficiency from the one-step and two-step RT-RPA was compared by using *E* gene as the detection target. The two-step RT-RPA was conducted using the optimized RT-RPA mixtures as described above, while the one-step RT-RPA was performed by adding all the reaction reagents at once and incubated at 37 °C for 30 min. The 50 μl-reaction mixture of the one-step RT-RPA contains 29.5 μl rehydration buffer, 10 μl RNA template, one freeze-dried Twist RPA reaction pellet, 2.5 μM RT-RPA forward and reverse primer mix, 0.4 mM dNTP mix, 200 U RevertAid reverse transcriptase, 6.6 μM T4 gene 32 protein, 100 U RNase H, 10 U RNase inhibitor, and 14 mM MgOAc. The RT-RPA products were loaded into 8% PAGE gel and the relative greyness of sample bands was quantified using ImageJ.

### Validation of RRCd in detection and discrimination of clinical samples

To evaluate the performance of RRCd on SARS-CoV-2 clinical samples, a total of 204 swabs were collected at SITHC from March 3^rd^ and to October 29^th^ 2021 and were used to perform the RRCd assay. Both lateral-flow and fluorescence-based readouts were recorded. A total of 96 clinical samples were used to detect SARS-CoV-2 by targeting *E* gene and *S* gene, and a total of 108 clinical samples were used to detect and discriminate the SARS-CoV-2 VOCs by targeting *S* gene 501 or 478. The RRCd results were analyzed and compared with the results of RT-qPCR and genome sequencing.

### RRCd using the directly released RNA

Viral RNAs from SARS-CoV-2 pseudovirus and clinical samples were released directly by a Sample release reagent kit (Sansure Biotech, Changsha, China) according to the manufacturer’s instructions with minor modifications. The swab storage solution and the sample release reagent were mixed in a ratio of 1:1 and incubated at 37 °C for 5 min before subsequent assays.

### Statistical analysis

Data were represented as mean ± S.D. from three independent experiments otherwise indicated. Statistical significances were analyzed by using the R (version 4.1.1). Unpaired one-tailed *t*-test was applied to investigate the differences between groups assuming *P* value < 0.05 as significant.

## Data Availability

All data produced in the present work are contained in the manuscript.

## Acknowledgements

We are grateful to Prof. Xian-En Zhang for generously providing the pseudovirus, and Prof. Hang Dai and Dr. Xiaonan Yang for constructive discussion on this project. This study was supported by the National Key R&D Program of China (2019YFA0904003, 2020YFA0909100), the Strategic Priority Research Program of the Chinese Academy of Sciences, China (XDB38020300), the Guangdong Basic and Applied Basic Research Foundation (2021A1515012511), and the General Administration of Customs Project (2020HK003).

## Author contributions

G.P.Z., Z.G.T. and W.Z. conceived and designed the study. G.T., Z.Z., W.T. and F.L. performed the experiments and analyzed data. All authors contributed to writing and editing of the manuscript. W.Z. supervised the research.

