## Supplementary Information for "RT-RPA-Cas12a-based discrimination of SARS-CoV-2 variants of concern"

[#]*These authors contributed equally.*

### Supplementary Figures

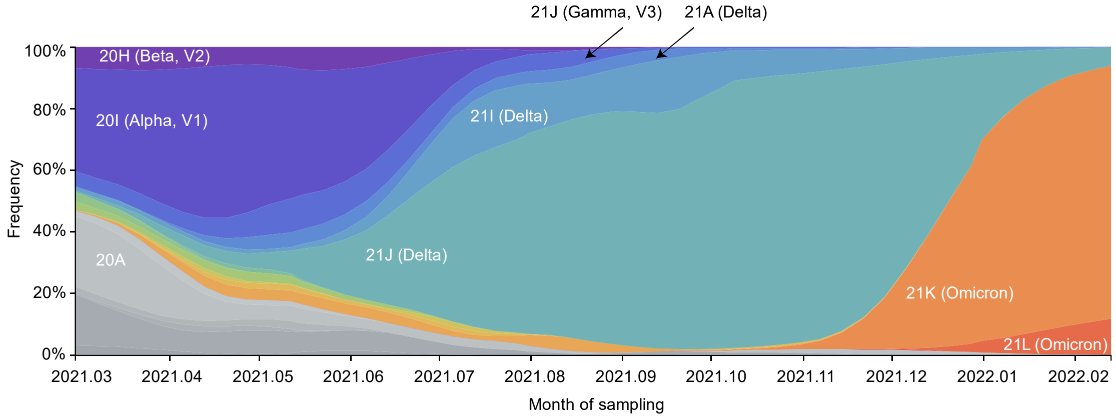

**Supplementary Figure 1. Global distribution of SARS-CoV-2 clades during the COVID-19 pandemic within the last 12 months.** Data were downloaded and adapted from GISAID (Shu and McCauley 2017) on February 21th 2022, filtered and processed using Nextstrain pipeline (Hadfield et al. 2018).

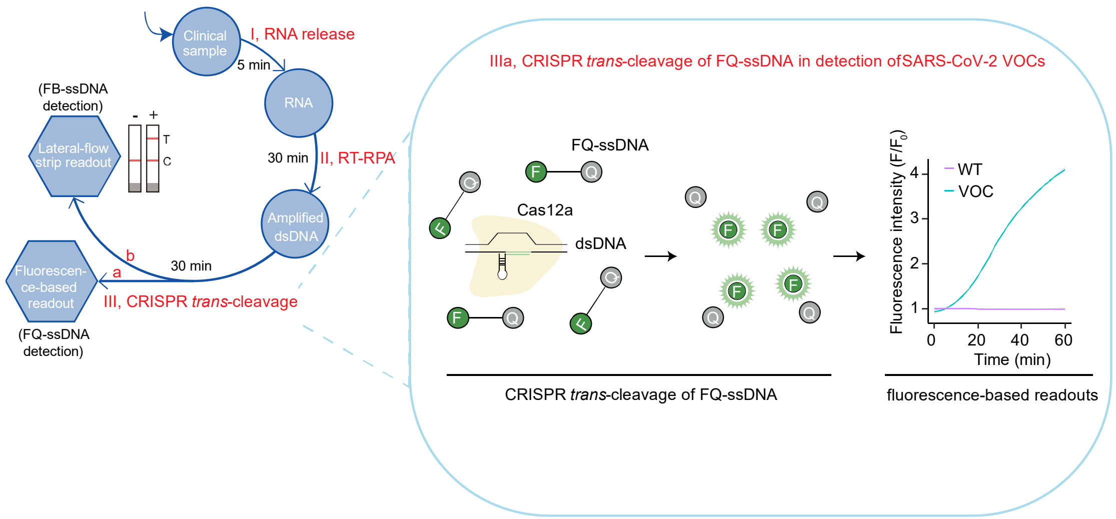

**Supplementary Figure 2. Schematics of the RRCd in discrimination of SARS-CoV-2 VOC using fluorescence-based readouts**. The analysis of fluorescence-based readouts is highlighted on the right panel. The dsDNA derived from SARS-CoV-2 VOCs is targeted and detected by crRNA-VOC and crRNA-WT, leading to the positive and negative test results, respectively. The relative fluorescence intensity ($F/F_0$) is calculated as the fluorescence signal versus the starting signal. FQ, fluorophore (FAM)-quencher (BHQ1); FB, FITC-Biotin.

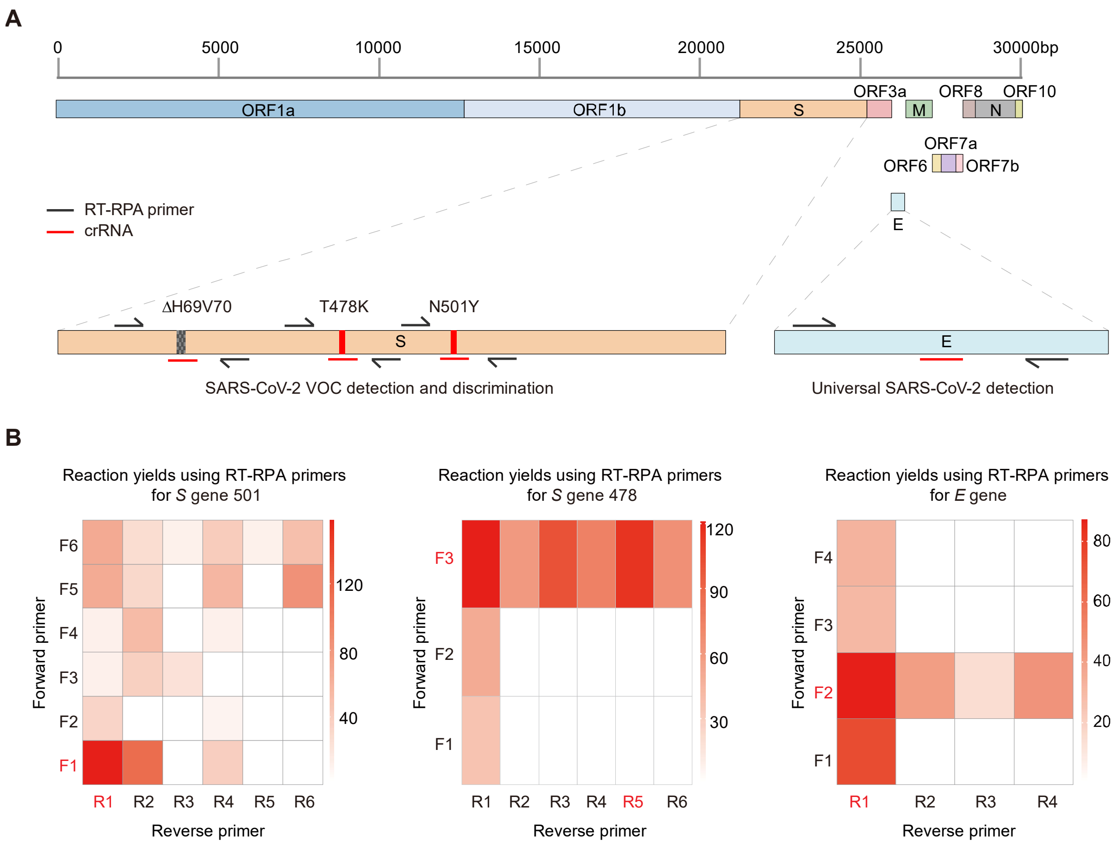

**Supplementary Figure 3. Systematic design and screening of optimal RT-RPA primers.** **A.** Schematic of SARS-CoV-2 genome (upper panel). The SARS-CoV-2 genome comprises of approximately 29,903 nucleotides and encodes 12 open reading frames, including the structural proteins of spike (S), nucleocapsid (N), membrane (M), and envelope (E). Among them, *S* and *E* genes were selected as the detection targets (lower panel). *E* gene was used for universal detection of SARS-CoV-2, while *S* gene was used for the detection and discrimination of SARS-CoV-2 VOCs. Forward and reverse RT-RPA primers (black arrows) and crRNA binding sites (red lines) for viral detection are indicated. **B.** Heatmap visualization for screening the optimal RT-RPA primers. The dsDNA derived from SARS-CoV-2 pseudovirus was amplified by RT-RPA using forward and reverse primers specific for *S* gene 501, *S* gene 478, and *E*

gene regions, respectively. The yields of each reaction were determined by agarose electrophoresis and quantified using ImageJ. The corresponding results were plotted as heatmaps by the R program (version 4.1.1). The selected optimal RT-RPA primers for the subsequent experiments are indicated in red.

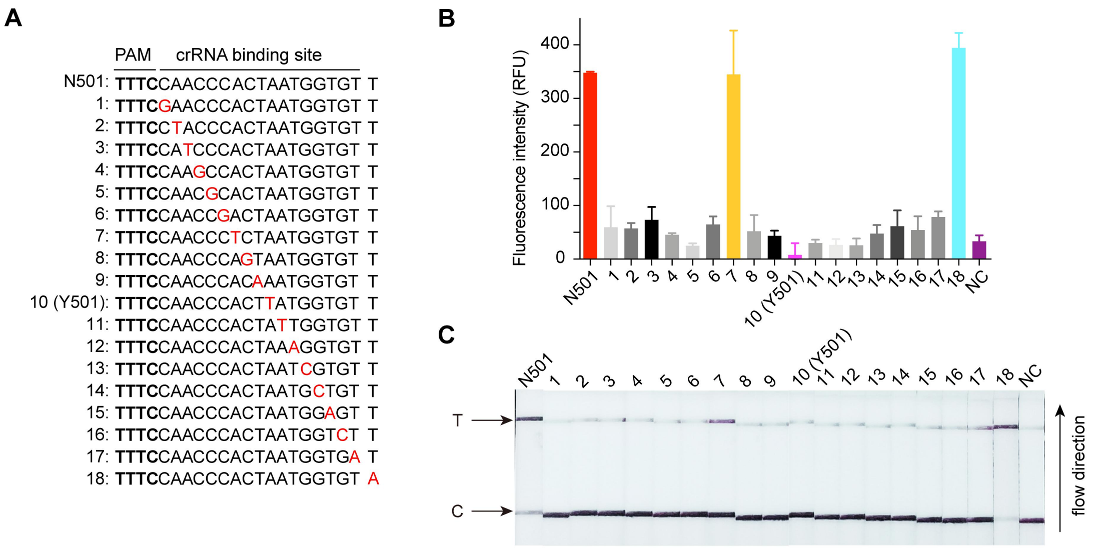

**Supplementary Figure 4. The specificity analysis of RRCd with crRNA-N501 targeting to the single-base mismatched *S* gene templates. A.** Representation of the wild-type (N501) and the single-base mismatched *S* gene templates. The PAM sequence (TTTC) is shown in bold and the mismatched crRNA binding site is shown in red. **B.** The signal-to-noise analysis of RRCd with fluorescence-based readouts. The results show that the RRCd has a high specificity in detection of *S* gene N501 except the mismatched nucleotide at the 7th of crRNA binding site. The *S* gene template with a mismatched nucleotide out of the crRNA binding site (18th) was used as a positive control. Data are presented as mean ± S.D. from three technical replicates. **C.** The signal-to-noise analysis of RRCd with lateral-flow-based readouts. The results are consistent with the fluorescence-based readouts in panel B. NC, negative control (RNase-free water).

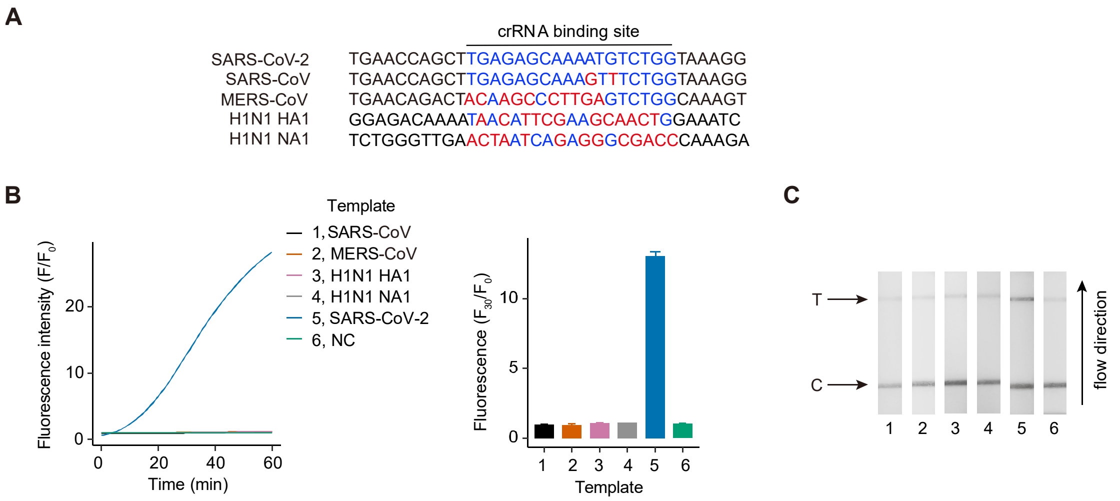

**Supplementary Figure 5. RRCd discriminates SARS-CoV-2 from other phylogenetic related viruses.A.** Sequence alignment of the *N* gene regions of SARS-CoV-2, SARS-CoV, MERS-CoV, H1N1 HA1 and H1N1 NA1. The crRNA binding site of SARS-CoV-2 *N* gene is indicated in blue, while the mismatched nucleotides in the *N* gene regions of other viruses are highlighted in red. **B-C.** Specificity analysis of RRCd in detecting *N* genes using SARS-CoV-2 crRNA-*N*. The results of fluorescence-based readouts (B) and lateral-flow readouts (C) are shown, respectively. The relative fluorescence intensity ($F/F_0$) is calculated as the fluorescence signal versus its starting signal. The fluorescence signals at the 30 min-reactions were quantified (middle panel) for each sample. NC, negative control (RNase-free water).

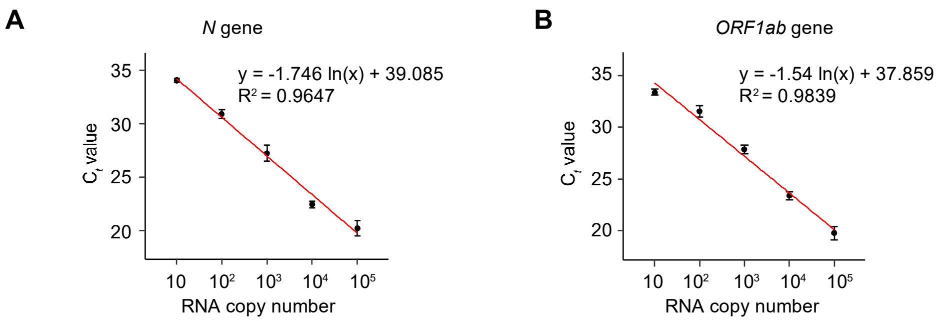

| RNA dilution (copies per reaction) | Mean ± S.D. $C_t$ (*N* gene) |
|---|---|
| $10^5$ | 20.23 ± 1.02 |
| $10^4$ | 22.47 ± 0.43 |
| $10^3$ | 27.26 ± 1.07 |
| $10^2$ | 30.95 ± 0.58 |
| 10 | 34.07 ± 0.25 |
| 0 | Undetectable |

| RNA dilution (copies per reaction) | Mean ± S.D. $C_t$ (*ORF1ab* gene) |
|---|---|
| $10^5$ | 19.79 ± 0.93 |
| $10^4$ | 23.40 ± 0.57 |
| $10^3$ | 27.89 ± 0.60 |
| $10^2$ | 31.57 ± 0.75 |
| 10 | 33.43 ± 0.39 |
| 0 | Undetectable |

**Supplementary Figure 6. Standard curve of $C_t$ values versus SARS-CoV-2 RNA copy numbers.** Serial dilutions of RNA extracted from SARS-CoV-2 pseudovirus were prepared and their corresponding $C_t$ values were determined by RT-qPCR. Standard curves for *N* (**A**) and *ORF1ab* (**B**) were generated by plotting RT-qPCR-derived $C_t$ values against the initial RNA copy numbers. Data are presented as mean ± S.D. from three technical replicates.

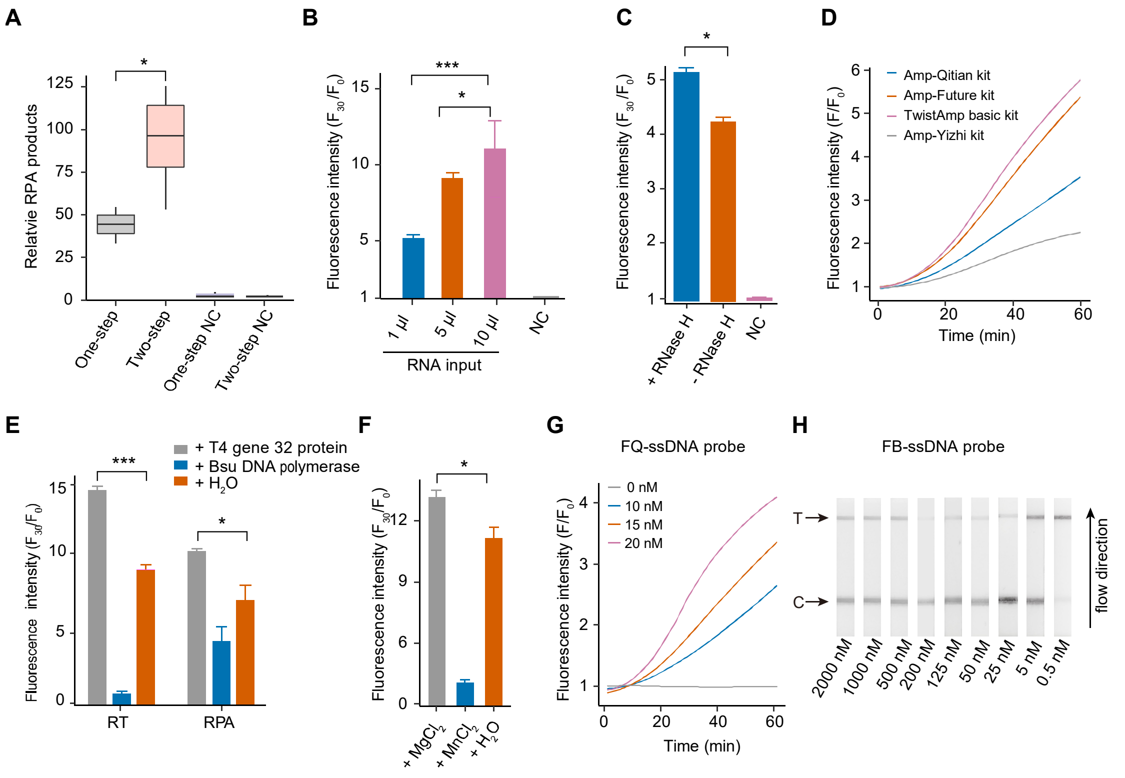

**Supplementary Figure 7. Systematic optimization of RRCd procedures.**

**A.** Comparison of the reaction yields between one-step and two-step RT-RPA. The results suggested that the two-step RT-RPA had a 47% higher amplification efficiency than that of the one-step RT-RPA. The RT-RPA products were quantified by analyzing the gel bands with ImageJ. Data are presented as mean ± S.D. from three independent experiment replicates.

**B.** Comparison of the fluorescence results using different RNA inputs in the RT reaction step. The input of 10 μl RNA displayed the highest fluorescence signal. The relative fluorescence intensity ($F/F_0$) is calculated as the fluorescence signal versus its starting signal. The fluorescence signals at the 30 min-reactions were quantified (middle panel) for each sample. Data are presented as mean ± S.D. from three technical replicates.

**C.** Comparison of the fluorescence results with and without RNase H in the RT reactions. As an endoribonuclease that specifically hydrolyzes the RNA-DNA hybrids,

RNase H was used to enhance the RT-RPA efficiency (Qian et al. 2020). Our results showed that the detection sensitivity of RRCd was increased with the adding of RNase H. Data are presented as mean ± S.D. from three technical replicates.

**D.** Comparison of the fluorescence results using different RPA kits in the RPA reaction step. The usage of TwistAmp Baic kit displayed the best fluorescence kinetic signal.

**E.** Comparison of the fluorescence results with and without T4 gene 32 protein (or Bsu DNA polymerase) in RT and RPA reactions. The results showed that the detection sensitivity of RRCd was increased by adding of T4 gene 32 protein in both RT and RPA reactions, but not of Bsu DNA polymerase. Data are presented as mean ± S.D. from three technical replicates.

**F**. Comparison of the fluorescence results between the usage of $MgCl_2$ and $MnCl_2$ in CRISPR-Cas12a cleavage reaction. The results showed that the detection sensitivity of RRCd was increased by $MgCl_2$ rather than $MnCl_2$. Data are presented as mean ± S.D. from three technical replicates.

**G.** Comparison of the fluorescence results using different concentrations of FQ-ssDNA probe in CRISPR-Cas12a cleavage reaction. The usage of 20 nM FQ-ssDNA probe displayed the highest fluorescence signal.

**H.** Comparison of the lateral-flow strip results using different concentrations of FB-ssDNA probe in CRISPR-Cas12a cleavage system without template. The usage of 200 nM FB-ssDNA probe displayed the lowest false-positive results.

*, $P < 0.05$; ***, $P < 0.01$. NC, negative control (RNase-free water).

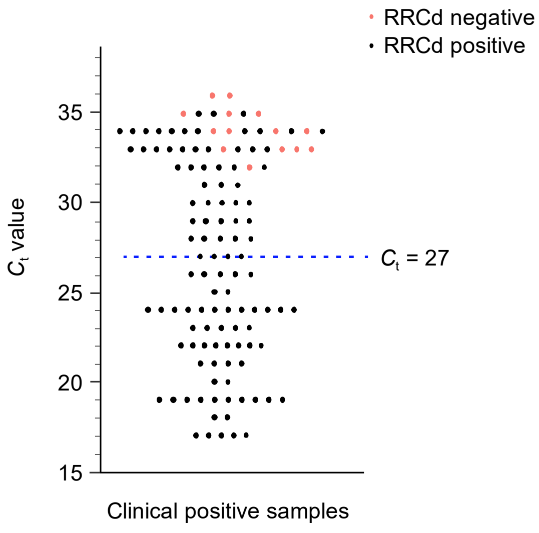

**Supplementary Figure 8. The distribution of $C_t$ values of SARS-CoV-2 clinical positive samples.** Scatter dot plot shows that the distribution of $C_t$ values of the 122 RT-qPCR-verified positive samples. The blue dotted line indicates a $C_t$ value of 27, corresponding to the mean $C_t$ value of these samples. Red dots, negative test results by RRCd; black dots, positive test results by RRCd.

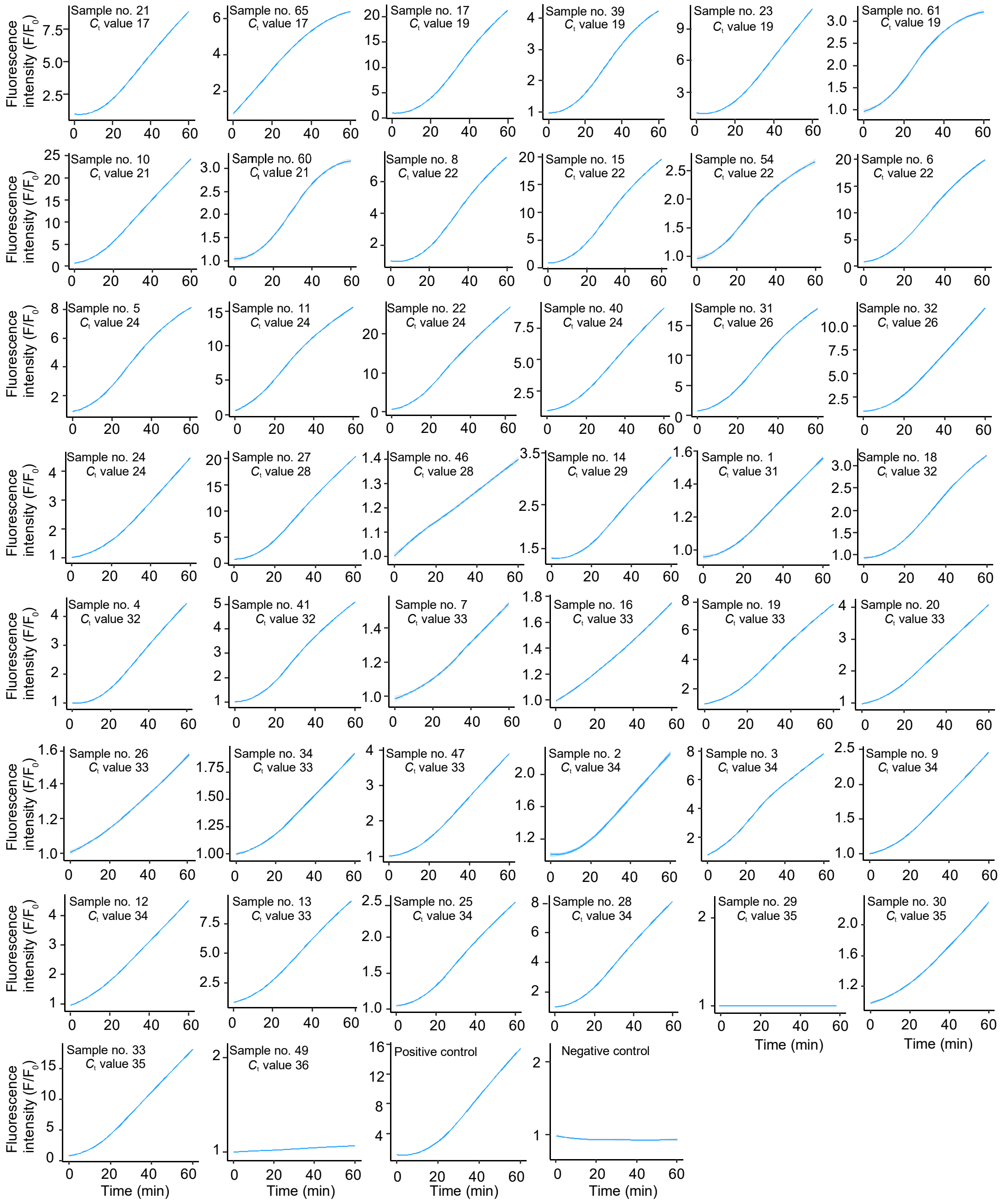

**Supplementary Figure 9. The detailed information of *E* gene detection by RRCd in clinical samples (Corresponding to Fig. 4).** The clinical sample was detected with crRNA-*E*, and the relative fluorescence intensities ($F/F_0$) over 60-min reactions for each positive sample are shown. RNase-free water was used as the negative control. The fitting curves were calculated using the LOESS method. The grey areas represent 95% confident intervals.

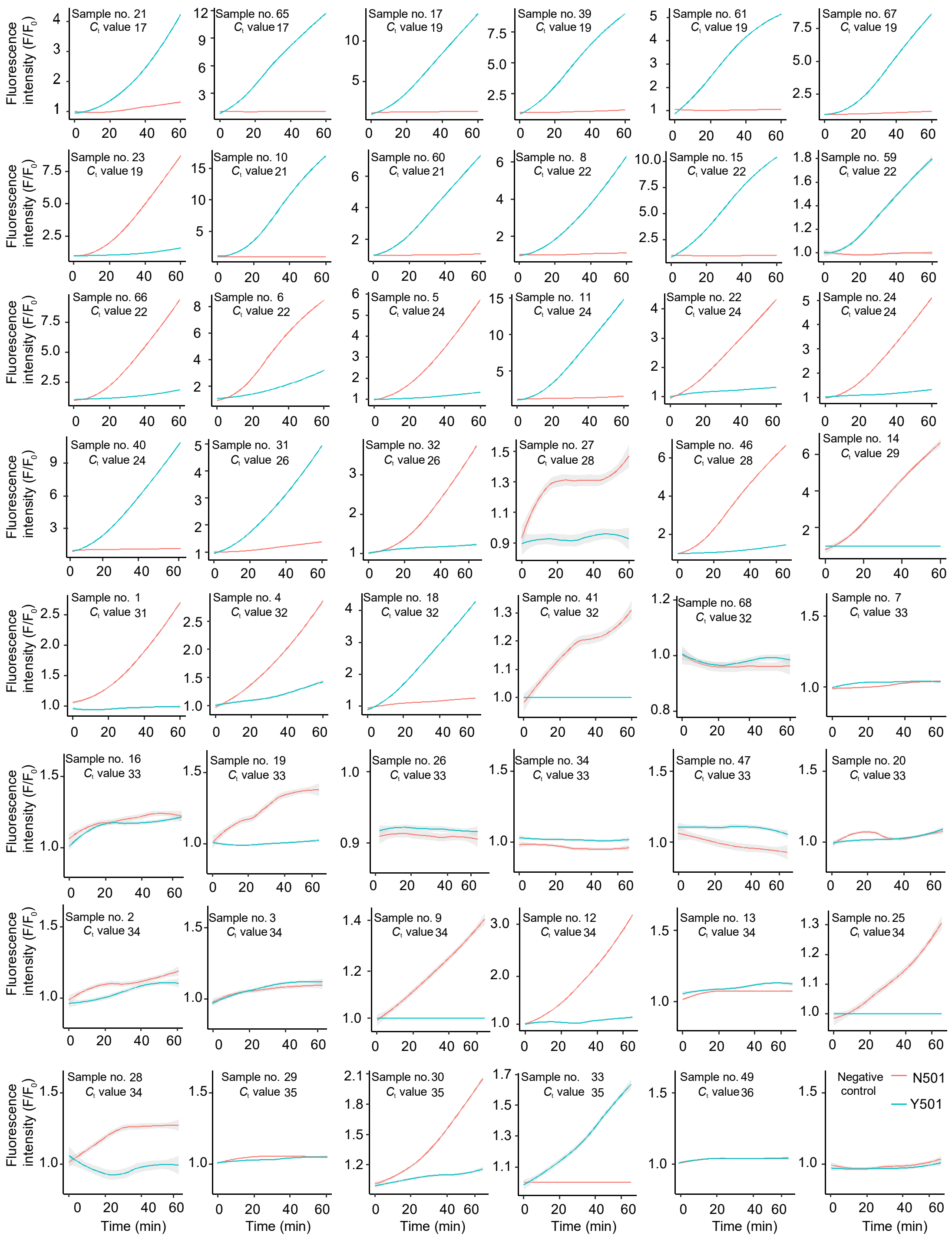

**Supplementary Figure 10. The detailed information of *S* gene 501 detection by RRCd in clinical samples (Corresponding to Fig. 5a).** The same clinical sample was detected with crRNA-N501 (red line) and crRNA-Y501 (sky blue line), respectively. The relative fluorescence intensities ($F/F_0$) over 60-min reactions for each positive sample are shown. RNase-free water was used as the negative control.

The fitting curves were calculated using the LOESS method. The grey areas represent 95% confident intervals.

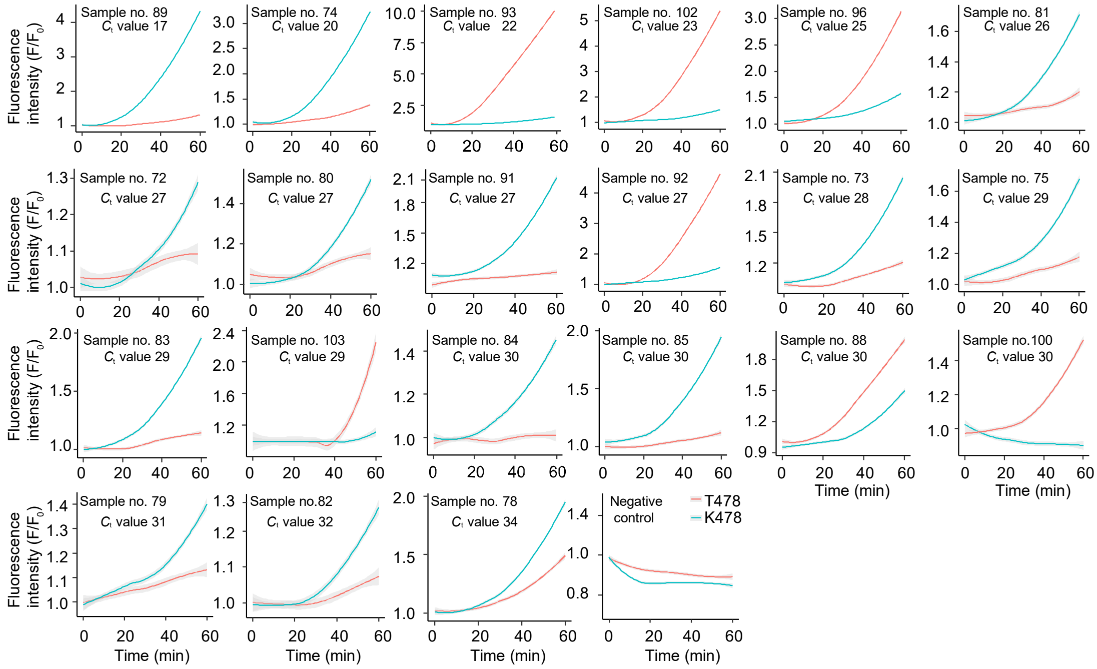

**Supplementary Figure S11. The detailed information of *S* gene 478 detection by RRCd in clinical samples (Corresponding to Fig. 5b).** The same clinical sample was detected with crRNA-T478 (red line) and crRNA-K478 (blue line), respectively. The relative fluorescence intensities ($F/F_0$) over 60-min reactions for each positive sample are shown. RNase-free water was used as the negative control. The fitting curves were calculated using the LOESS method. The grey areas represent 95% confident intervals.

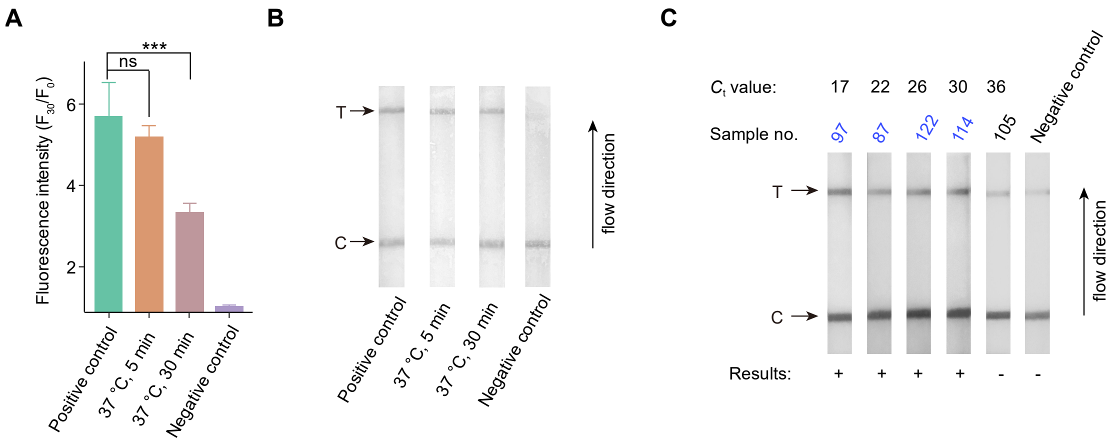

**Supplementary Figure 12. RRCd using the directly released sample**.

**A-B.** Fluorescence-based readouts (A) and lateral-flow readouts (B) of RRCd using the directly released pseudovirus RNA. The RNAs were released by a Sample release reagent kit (Sansure Biotech, Changsha, China) at 37 °C for 5 min or 30 min, without extraction and purification. *E* gene was used as the detection target and the results of corresponding fluorescence intensities ($F_{30}/F_0$) were monitored. RRCd showed a comparable detection sensitivity when using the 5-min treated sample. The extract from the same volume of pseudovirus was used as the positive control, and the RNase-free water was used as the negative control. Data are represented as mean ± S.D. from three independent experiments. ***, $P < 0.01$; ns, $P \geq 0.05$. **c.** Lateral-flow readout of RRCd for the detection of SARS-CoV-2 *E* gene using the directly released clinical samples. +, positive; －, negative.

**Supplementary Table 1. Primers, crRNAs and probes used in this study.**

| Name | Sequence (5’-3’) | Target or purpose | Source |
|---|---|---|---|
| **RT-RPA primer** | | | |
| ERPAF1 | tactcattcgtttcggaagagacaggtacgtt | SARS-CoV-2 *E* gene | This study |
| ERPAR1 | cagatttttaacacgagagtaaacgtaaaaagaa | SARS-CoV-2 *E* gene | This study |
| ERPAF2 | gaagagacaggtacgttaatagttaatagc | SARS-CoV-2 *E* gene | This study |
| ERPAR2 | gaaggttttacaagactcacgttaacaatatt | SARS-CoV-2 *E* gene | This study |
| ERPAF3 | acgttaatagttaatagcgtacttctttttct | SARS-CoV-2 *E* gene | This study |
| ERPAR3 | ctagaagaattcagatttttaacacgagagta | SARS-CoV-2 *E* gene | This study |
| ERPAF4 | ttgctttcgtggtattcttgctagttacacta | SARS-CoV-2 *E* gene | This study |
| ERPAR4 | aaagaaggttttacaagactcacgttaacaat | SARS-CoV-2 *E* gene | This study |
| S501RPAF1 | tgtatagattgtttaggaagtctaatctcaa | SARS-CoV-2 *S* gene | This study |
| S501RPAR1 | agactcagtaagaacacctgtgcctgttaa | SARS-CoV-2 *S* gene | This study |
| S501RPAF2 | taggaagtctaatctcaaacctttga | SARS-CoV-2 *S* gene | This study |
| S501RPAR2 | gtgcatgtagaagttcaaaagaaagtacta | SARS-CoV-2 *S* gene | This study |
| S501RPAF3 | cttgtaatggtgttgaaggttttaattgt | SARS-CoV-2 *S* gene | This study |
| S501RPAR3 | ctgtgcctgttaaaccattgaagttgaaat | SARS-CoV-2 *S* gene | This study |
| S501RPAF4 | atggtgttgaaggttttaattgttactttc | SARS-CoV-2 *S* gene | This study |
| S501RPAR4 | taaaccattgaagttgaaattgacacattt | SARS-CoV-2 *S* gene | This study |
| S501RPAF5 | gttactttcctttacaatcatatggtttcc | SARS-CoV-2 *S* gene | This study |
| S501RPAR5 | cctgttaaaccattgaagttgaaattga | SARS-CoV-2 *S* gene | This study |
| S501RPAF6 | ccttgtaatggtgttgaaggttttaattgt | SARS-CoV-2 *S* gene | This study |
| S501RPAR6 | tctgtatggttggtaaccaacaccattagt | SARS-CoV-2 *S* gene | This study |
| NRPAF | gttcctcatcacgtagtcgcaacagttcaagaa | SARS-CoV-2 *N* gene | This study |
| NRPAR | gcttcttagaagcctcagcagcagatttcttag | SARS-CoV-2 *N* gene | This study |
| S478RPAF1 | cttgattctaaggttggtggtaattataatttac | SARS-CoV-2 *S* gene | This study |
| S478RPAF2 | cttgattctaaggttggtggtaatttataattac | SARS-CoV-2 *S* gene | This study |
| S478RPAF3 | accagatgattttacaggctgcgttatagcttg | SARS-CoV-2 *S* gene | This study |
| S478RPAR1 | gtaacaattaaaaccttcaacaccatttacaagg | SARS-CoV-2 *S* gene | This study |
| S478RPAR2 | gttggtaaccaacaccataagtgggttggaaacc | SARS-CoV-2 *S* gene | This study |
| S478RPAR3 | ttgtaaaggaaagtaacaattaaaaccttcaacac catttacaagg | SARS-CoV-2 *S* gene | This study |
| S478RPAR4 | gtaacaattaaaacctttaacaccattacaagg | SARS-CoV-2 *S* gene | This study |
| S478RPAR5 | caattaaaacctttaacaccattacaa | SARS-CoV-2 *S* gene | This study |
| S478RPAR6 | agtaacaattaaaacctttaacacc | SARS-CoV-2 *S* gene | This study |
| S6970RPAF | tttccaatgttacttggttccatgtttactat | SARS-CoV-2 *S* gene | This study |
| S6970RPAR | ttaacaataagtagggactgggtcttcgaatc | SARS-CoV-2 *S* gene | This study |
| SARS-CoV NRPAF | aatggctagcggaggtggtgaaactgccctcgc | SARS-CoV *N* gene | This study |
| SARS-CoV NRPAR | aatgcagaggcacttggagcaaattgtgcaattt gcggc | SARS-CoV *N* gene | This study |
| MERS-CoV NRPAF | aaggttcaagatcaggaaactctacccgcggc | MERS-CoV *N* gene | This study |

| | | | |
|---|---|---|---|
| MERS-CoV NRPAR | actggctgtaggagcaagctcagcaatttggggc | MERS-CoV *N* gene | This study |
| HA1NRPAF | aatgaactattactggacactagtagagccgg | H1N1 HA1 *N* gene | This study |
| HA1RPAR | tggccccaaataggcctctagattgaatagacggg | H1N1 HA1 *N* gene | This study |
| NA1RPAF | aaacggttttgagatgatttgggatccgaacgg | H1N1 HN1 *N* gene | This study |
| NA1RPAR | tggtaaatggcaactcagcaccgtctggcc | H1N1 HN1 *N* gene | This study |
| **PCR primer** | | | |
| M13-Forward | cccagtcacgacgttgtaaaacg | *In vitro* transcription | This study |
| M13-Reverse | agcggataacaatttcacacagg | *In vitro* transcription | This study |
| SN501-1-F | aatcatatggtttcgaacccactaatggtg | Construction of SARS-CoV-2 *S* gene 501 variant-1 | This study |
| SN501-1-R | caccattagtgggttcgaaaccatatgatt | | This study |
| SN501-2-F | aatcatatggtttcctacccactaatggtg | Construction of SARS-CoV-2 *S* gene 501 variant-2 | This study |
| SN501-2-R | caccattagtgggtaggaaaccatatgatt | | This study |
| SN501-3-F | atcatatggtttccatcccactaatggtgt | Construction of SARS-CoV-2 *S* gene 501 variant-3 | This study |
| SN501-3-R | acaccattagtgggatggaaaccatatgat | | This study |
| SN501-4-F | atcatatggtttccaagccactaatggtgtt | Construction of SARS-CoV-2 *S* gene 501 variant-4 | This study |
| SN501-4-R | aacaccattagtggcttggaaaccatatgat | | This study |
| SN501-5-F | catatggtttccaacgcactaatggtgttg | Construction of SARS-CoV-2 *S* gene 501 variant-5 | This study |
| SN501-5-R | caacaccattagtgcgttggaaaccatatg | | This study |
| SN501-6-F | atatggtttccaaccgactaatggtgttgg | Construction of SARS-CoV-2 *S* gene 501 variant-6 | This study |
| SN501-6-R | ccaacaccattagtcggttggaaaccatat | | This study |
| SN501-7-F | tatggtttccaaccctctaatggtgttggt | Construction of SARS-CoV-2 *S* gene 501 variant-7 | This study |
| SN501-7-R | accaacaccattagagggttggaaaccata | | This study |
| SN501-8-F | atggtttccaacccagtaatggtgttggtt | Construction of SARS-CoV-2 *S* gene 501 variant-8 | This study |
| SN501-8-R | aaccaacaccattactgggttggaaaccat | | This study |
| SN501-9-F | tggtttccaacccacaaatggtgttggtta | Construction of SARS-CoV-2 *S* gene 501 variant-9 | This study |
| SN501-9-R | taaccaacaccatttgtgggttggaaacca | | This study |
| SN501-10-F | ggtttccaacccacttatggtgttggttac | Construction of SARS-CoV-2 *S* gene 501 variant-10 | This study |
| SN501-10-R | gtaaccaacaccataagtgggttggaaacc | | This study |
| SN501-11-F | gtttccaacccactattggtgttggttacc | Construction of SARS-CoV-2 *S* gene 501 variant-11 | This study |
| SN501-11-R | ggtaaccaacaccaatagtgggttggaaac | | This study |
| SN501-12-F | gtttccaacccactaaaggtgttggttacca | Construction of SARS-CoV-2 *S* gene 501 variant-12 | This study |
| SN501-12-R | tggtaaccaacacctttagtgggttggaaac | | This study |
| SN501-13-F | gtttccaacccactaatcgtgttggttaccaa | Construction of | This study |

| | | | |
|---|---|---|---|
| SN501-13-R | ttggtaaccaacacgattagtgggttggaaac | SARS-CoV-2 *S* gene 501 variant-13 | This study |
| SN501-14-F | tccaacccactaatgctgttggttaccaac | Construction of SARS-CoV-2 *S* gene 501 variant-14 | This study |
| SN501-14-R | gttggtaaccaacagcattagtgggttgga | | This study |
| SN501-15-F | tccaacccactaatggagttggttaccaacc | Construction of SARS-CoV-2 *S* gene 501 variant-15 | This study |
| SN501-15-R | ggttggtaaccaactccattagtgggttgga | | This study |
| SN501-16-F | aacccactaatggtcttggttaccaaccata | Construction of SARS-CoV-2 *S* gene 501 variant-16 | This study |
| SN501-16-R | tatggttggtaaccaagaccattagtgggtt | | This study |
| SN501-17-F | aacccactaatggtgatggttaccaaccat | Construction of SARS-CoV-2 *S* gene 501 variant-17 | This study |
| SN501-17-R | atggttggtaaccatcaccattagtgggtt | | This study |
| SN501-18-F | aacccactaatggtgtaggttaccaaccat | Construction of SARS-CoV-2 *S* gene 501 variant -18 | This study |
| SN501-18-R | atggttggtaacctacaccattagtgggtt | | This study |
| **crRNA** | | | |
| crRNA-*E* | AAUUUCUACUGUUGUAGAUc aagacucacguuaacaa | SARS-CoV-2 *E* gene | This study |
| crRNA-N501 | AAUUUCUACUGUUGUAGAUc aacccacuaauggugu | SARS-CoV-2 *S* gene | This study |
| crRNA-Y501 | AAUUUCUACUGUUGUAGAUc aacccacuuauggugu | SARS-CoV-2 *S* gene | This study |
| SARS-CoV-2 crRNA-*N* | AAUUUCUACUGUUGUAGAUc cagacauuuugcucuca | SARS-CoV-2 *N* gene | This study |
| SARS-CoV - crRNA-*N* | AAUUUCUACUGUUGUAGAUc cagaaacuuugcucuca | SARS-CoV *N* gene | This study |
| MERS-CoV - crRNA-*N* | AAUUUCUACUGUUGUAGAUc cagacucaagggcuugu | MERS-CoV *N* gene | This study |
| H1N1 HA1- crRNA-*N* | AAUUUCUACUGUUGUAGAUc aguugcuucgaauguua | H1N1 NA1 *N* gene | This study |
| H1N1 NA1- crRNA-*N* | AAUUUCUACUGUUGUAGAUg gucgccccucugauuagu | H1N1 NA1 *N* gene | This study |
| crRNA-H69-V70 | AAUUUCUACUGUUGUAGAUc uauacaugucucuggg | SARS-CoV-2 *S* gene | This study |
| crRNA-ΔH69-V70 | AAUUUCUACUGUUGUAGAUc uaucucugggaccaau | SARS-CoV-2 *S* gene | This study |
| crRNA-T478 | AAUUUCUACUGUUGUAGAUu gugguaauguuccaca | SARS-CoV-2 *S* gene | This study |
| crRNA- K478 | AAUUUCUACUGUUGUAGAUu gugguaauguuccaaa | SARS-CoV-2 *S* gene | This study |
| **Probe** | | | |
| FQ-ssDNA | FAM/gctaatcg/BHQ1/ | fluorescence-based readout | Sangon |
| FB-ssDNA | /FITC/gattagcgtacgcacgttac/Biotin/ | lateral-flow readout | Zoonbio Biotechnology |

**Supplementary Table 2. The synthetic DNA sequences in this study.**

| Name | DNA sequence |
|---|---|
| T7 promoter | TAATACGACTCACTATAGG |
| SARS-CoV-2 *S* gene N501 | TGTATAGATTGTTTAGGAAGTCTAATCTCAAACCTTTTGAG<br>AGAGATATTTCAACTGAAATCTATCAGGCCGGTAGCACACC<br>TTGTAATGGTGTTGAAGGTTTTAATTGTTACTTTCCTTTACA<br>ATCATATGGTTTCCAACCCACTAATGGTGTTGGTTACCAAC<br>CATACAGAGTAGTAGTACTTTCTTTTGAACTTCTACATGCA<br>CCAGCAACTGTTTGTGGACCTAAAAAGTCTACTAATTTGGT<br>TAAAAACAAATGTGTCAATTTCAACTTCAATGGTTTAACAG<br>GCACAGGTGTTCTTACTGAGTCTAACAAAAAGTTTCTGCC<br>TTTCCAACAATTTGGCAGAGACATTGCTGACACTACTGATG<br>CTGTCCGTGATCCACAGACACTTGAGATTCTTGACATTACA<br>CCATGTTCTTTTGGTGGTGTCAGTGTTATAACACCAGGAAC<br>AAATACTTCTAACCAGGTTGCTGTTCTTTATCAGGATGTTA<br>ACTGCACAGAAGTCCCTGTTGCTATTCATGCAGATCAACTT<br>ACTCCTACTTGGCGTGTTTATTCTACAGGTTCTAATGTTTTT<br>CAAACACGTGCAGGCTGTTTAATAGGGGCTGAACATGTCA<br>ACAACTCATATGAGTGTGACATACCCATTGGTGCAGGTATA<br>TGCGCT |
| SARS-CoV *N* gene | GATCACTAATACGACTCACTATAGGGCATCACGTAGTCGCG<br>GTAATTCAAGAAATTCAACTCCTGGCAGCAGTAGGGGAAA<br>TTCTCCTGCTCGAATGGCTAGCGGAGGTGGTGAAACTGCC<br>CTCGCGCTATTGCTGCTAGACAGATTGAACCAGCTTGAGA<br>GCAAAGTTTCTGGTAAAGGCCAACAACAACAAGGCCAAA<br>CTGTCACTAAGAAATCTGCTGCTGAGGCATCTAAAAAGCC<br>TCGCCAAAAACGTACTGCCACAAAACAGTACAACGTCACT<br>CAAGCATTTGGGAGACGTGGTCCAGAACAAACCCAAGGA<br>AATTTCGGGGACCAAGACCTAATCAGACAAGGAACTGATT<br>ACAAACATTGGCCGCAAATTGCACAATTTGCTCCAAGTGC<br>CTCTGCATTCTTTGGAATGTCACGCAT |
| MERS-CoV *N* gene | GATCACTAATACGACTCACTATAGGGGCAGAAACTCTTCCA<br>GATCTAGTTCACAAGGTTCAAGATCAGGAAACTCTACCCG<br>CGGCACTTCTCCAGGTCCATCTGGAATCGGAGCAGTAGGA<br>GGTGATCTACTTTACCTTGATCTTCTGAACAGACTACAAGC<br>CCTTGAGTCTGGCAAAGTAAAGCAATCGCAGCCAAAAGTA<br>ATCACTAAGAAAGATGCTGCTGCTGCTAAAAATAAGATGC<br>GCCACAAGCGCACTTCCACCAAAAGTTTCAACATGGTGCA<br>AGCTTTTGGTCTTCGCGGACCAGGAGACCTCCAGGGAAAC<br>TTTGGTGATCTTCAATTGAATAAACTCGGCACTGAGGACCC<br>ACGTTGGCCCCAAATTGCTGAGCTTGCTCCTACAGCCAGT<br>GCTTTTATGGGTATGTCGCAATT |
| H1N1 HA1 *N* gene | GATCACTAATACGACTCACTATAGGGTCAAGATACAGCAAG<br>AAGTTCAAGCCGGAAATAGCAATAAGACCCAAAGTGAGG<br>GATCAAGAAGGGAGAATGAACTATTACTGGACACTAGTAG<br>AGCCGGGAGACAAAATAACATTCGAAGCAACTGGAAATCT<br>AGTGGTACCGAGATATGCATTCGCAATGGAAAGAAATGCT<br>GGATCTGGTATTATCATTTCAGATACACCAGTCCACGATTGC |

| | |
|---|---|
| | AATACAACTTGTCAAACACCCAAGGGTGCTATAAACACCA GCCTCCCATTTCAGAATATACATCCGATCACAATTGGAAAA TGTCCAAAATATGTAAAAAGCACAAAATTGAGACTGGCCA CAGGATTGAGGAATATCCCGTCTATTCAATCTAGAGGCCTA TTTGGGGCCATTGCCGGTTTCA |
| H1N1 NA1 *N* gene | GATCACTAATACGACTCACTATAGGGAGTATCGTCTAATGG AGCAAATGGAGTAAAAGGGTTTTCATTCAAATACGGCAAT GGTGTTTGGATAGGGAGAACTAAAAGCATTAGTTCAAGAA ACGGTTTTGAGATGATTTGGGATCCGAACGGATGGACTGG GACAGACAATAACTTCTCAATAAAGCAAGATATCGTAGGA ATAAATGAGTGGTCAGGATATAGCGGGAGTTTTGTTCAGCA TCCAGAACTAACAGGGCTGGATTGTATAAGACCTTGCTTCT GGGTTGAACTAATCAGAGGGCGACCCAAAGAGAACACAA TCTGGACTAGCGGGAGCAGCATATCCTTTTGTGGTGTAAAC AGTGACACTGTGGGTTGGTCTTGGCCAGACGGTGCTGAGT TGCCATTTACCATTGACAAGTAA |

172

**Supplementary Table 3. SARS-CoV-2 pseudovirus used in this study.**

| | |
|---|---|
| SARS-CoV-2 ab II EMNS S- MT | *S* gene (NC_045512.2) selected 22517-23866 |
| | AGAGTCCAACCAACAGAATCTATTGTTAGATTTCCTAATATTACAAACTTGTGCCCTTTTGGTGAAGTTTTAACGCCACCAGATTTGCATCTGTTTATGCTTGGAACAGGAAGAGAATCAGCAACTGTGTTGCTGATTATTCTGTCCTATATAATTCCGCATCATTTTCCACTTTTAAGTGTTATGGAGTGTCTCCTACTAAATTAAATGATCTCTGCTTTACTAATGTCTATGCAGATTCATTTGTAATTAGAGGTGATGAAGTCAGACAAATCGCTCCAGGGCAAACTGGAAAGATTGCTGATTATAATTATAAATTACCAGATGATTTTACAGGCTGCGTTATAGCTTGGAATTCTAAA(N439K)AATCTTGATTCTAAGGTTGGTGGTAATTATAATTACCTGTATAGATTGTTTAGGAAGTCTAATCTCAAACCTTTTGAGAGAGATATTTCAACTGAAATCTATCAGGCCGGTAGCACACCTTGTAATGGTGTTGAAGGTTTTAATTGTTACTTTCCTTTACAATCATATGGTTTCCAACCCACTT(N501Y)ATGGTGTTGGTTACCAACCATACAGAGTAGTAGTACTTTCTTTTGAACTTCTACATGCACCAGCAACTGTTTGTGGACCTAAAAAGTCTACTAATTTGGTTAAAAACAAATGTGTCAATTTCAACTTCAATGGTTTAACAGGCACAGGTGTTCTTACTGAGTCTAACAAAAAGTTTCTGCCTTTCCAACAATTTGGCAGAGACATTGA(A570D)TGACACTACTGATGCTGTCCGTGATCCACAGACACTTGAGATTCTTGACATTACACCATGTTCTTTTGGTGGTGTCAGTGTTATAACACCAGGAACAAATACTTCTAACCAGGTTGCTGTTCTTTATCAGGG(D614G)TGTTAACTGCACAGAAGTCCCTGTTGCTATTCATGCAGATCAACTTACTCCTACTTGGCGTGTTTATTCTACAGGTTCTAATGTTTTTCAAACACGTGCAGGCTGTTTAATAGGGGCTGAACATGTCAACAACTCATATGAGTGTGACATACCCATTGGTGCAGGTATATGCGCTAGTTATCAGACTCAGACTAATTCTCA(P681H)TCGGCGGGCACGTAGTGTAGCTAGTCAATCCATCATTGCCTACACTATGTCACTTGGTGCAGAAAATTCAGTTGCTTACTCTAATAACTCTATTGCCATACCCAT(T716I)AAATTTTACTATTAGTGTTACCACAGAAATTCTACCAGTGTCTATGACCAAGACATCAGTAGATTGTACAATGTACATTTGTGGTGATTCAACTGAATGCAGCAATCTTTTGTTGCAATATGGCAGTTTTTGTACACAATTAAACCGTGCTTTAACT |
| SARS-COV-2-MT-B.1.617.2 | *S* gene (NC_045512.2) selected 22009-23814 |
| | CAAAAGTTGGATGGAAAGTG(E156G&Del 157F、168R)GAGTTTATTCTAGTGCGAATAATTGCACTTTTGAATATGTCTCTCAGCCTTTTCTTATGGACCTTGAAGGAAAACAGGGTAATTTCAAAAATCTTAGGGAATTTGTGTTTAAGAATATTGATGGTTATTTTAAAATATATTCTAAGCACACGCCTATTAATTTAGTGCGTGATCTCCCTCAGGGTTTTTCGGCTTTAGAACCATTGGTAGATTTGCCAATAGGTATTAACATCACTAGGTTTCAAACTTTACTTGCTTTACATAGAAGTTATTTGACTCCTGGTGATTCTTCTTCAGGTTGGACAGCTGGTGCTGCAGCTTATTATGTGGGTTATCTTCAACCTAGGACTTTTCTATTAAAA |

| | |
|---|---|
| | TATAATGAAAATGGAACCATTACAGATGCTGTAGACTGTG<br>CACTTGACCCTCTCTCAGAAACAAAGTGTACGTTGAAAT<br>CCTTCACTGTAGAAAAAGGAATCTATCAAACTTCTAACTT<br>TAGAGTCCAACCAACAGAATCTATTGTTAGATTTCCTAATA<br>TTACAAACTTGTGCCCTTTTGGTGAAGTTTTTAACGCCAC<br>CAGATTTGCATCTGTTTATGCTTGGAACAGGAAGAGAATC<br>AGCAACTGTGTTGCTGATTATTCTGTCCTATATAATTCCGC<br>ATCATTTTCCACTTTTAAGTGTTATGGAGTGTCTCCTACTA<br>AATTAAATGATCTCTGCTTTACTAATGTCTATGCAGATTCA<br>TTTGTAATTAGAGGTGATGAAGTCAGACAAATCGCTCCAG<br>GGCAAACTGGAAAGATTGCTGATTATAATTATAAATTACC<br>AGATGATTTTACAGGCTGCGTTATAGCTTGGAATTCTAAC<br>AATCTTGATTCTAAGGTTGGTGGTAATTATAATTACCG(L45<br>2R)GTATAGATTGTTTAGGAAGTCTAATCTCAAACCTTTTG<br>AGAGAGATATTTCAACTGAAATCTATCAGGCCGGTAGCAA<br>(T478K)ACCTTGTAATGGTGTTGAAGGTTTTAATTGTTACT<br>TTCCTTTACAATCATATGGTTTCCAACCCACTAATGGTGTT<br>GGTTACCAACCATACAGAGTAGTAGTACTTTCTTTTGAAC<br>TTCTACATGCACCAGCAACTGTTTGTGGACCTAAAAAGTC<br>TACTAATTTGGTTAAAAACAAATGTGTCAATTTCAACTTC<br>AATGGTTTAACAGGCACAGGTGTTCTTACTGAGTCTAACA<br>AAAAGTTTCTGCCTTTCCAACAATTTGGCAGAGACATTGC<br>TGACACTACTGATGCTGTCCGTGATCCACAGACACTTGA<br>GATTCTTGACATTACACCATGTTCTTTTGGTGGTGTCAGT<br>GTTATAACACCAGGAACAAATACTTCTAACCAGGTTGCTG<br>TTCTTTATCAGGG(D614G)TGTTAACTGCACAGAAGTCCCT<br>GTTGCTATTCATGCAGATCAACTTACTCCTACTTGGCGTGT<br>TTATTCTACAGGTTCTAATGTTTTTCAAACACGTGCAGGC<br>TGTTTAATAGGGGCTGAACATGTCAACAACTCATATGAGT<br>GTGACATACCCATTGGTGCAGGTATATGCGCTAGTTATCA<br>GACTCAGACTAATTCTCG(P681R)TCGGCGGGCACGTAGT<br>GTAGCTAGTCAATCCATCATTGCCTACACTATGTCACTTGG<br>TGCAGAAAATTCAGTTGCTTACTCTAATAACTCTATTGCCA<br>TACCCACAAATTTTACTATTAGTGTTACCACAGAAATTCTA<br>CCAGTGTCTATGACCAAGACATCAGTAGATTGTACAATGT<br>ACATTTGTGGTGATTCAACTGAATGCAGCAA |

The mutation sites in the pseudoviruses are indicated.
